# Polyol pathway dysregulation in CSF links glucose metabolism to tau pathology independently of amyloid and genetic predisposition

**DOI:** 10.64898/2026.05.06.26352559

**Authors:** Marc Clos-Garcia, Asger Wretlind, Tik Muk, Kourosh Hooshmand, Anja Hviid Simonsen, Laura M. Winchester, Petroula Proitsi, Riccardo E. Marioni, Tarunveer S. Ahluwalia, Thomas Kümler, Steen Gregers Hasselbalch, Cristina Legido-Quigley, the Alzheimer’s Disease Neuroimaging Initiative, the Alzheimer’s Disease Metabolomics Consortium

**Author notes:** Corresponding authors: Steen Gregers Hasselbalch, Rigshospitalet – Neurocentret, Blegdamsvej 3, København N, 2200.; Cristina Legido-Quigley, Franklin-Wilkins Building, 150 Stamford Street, London, SE1 9NH,34. Data used in preparation of this article were obtained from the Alzheimer’s Disease Neuroimaging Initiative (ADNI) database (adni.loni.usc.edu). As such, the investigators within the ADNI contributed to the design and implementation of ADNI and/or provided data but did not participate in the analysis or writing of this report. A complete listing of ADNI investigators can be found at: http://adni.loni.usc.edu/wp-content/uploads/how_to_apply/ADNI_Acknowledgement_List.pdf. Data used in preparation of this article were generated by the Alzheimer’s Disease Metabolomics Consortium (ADMC). As such, the investigators within the ADMC provided data but did not participate in the analysis or writing of this report. A complete listing of ADMC investigators can be found at: https://sites.duke.edu/adnimetab/team/.

## Abstract

Dementia affects approximately 60 million people worldwide, yet molecular mechanisms linking early neuropathological changes to clinical progression remain poorly understood. We performed targeted and untargeted metabolomics in plasma and cerebrospinal fluid (CSF) from 166 memory clinic patients spanning no cognitive impairment, mild cognitive impairment due to Alzheimer’s disease (AD), AD dementia, and mixed AD–cerebrovascular dementia.

Using a data-driven approach, we identified a CSF polyol signature characterized by elevated sorbitol, meso-erythritol, and d-glucose/erythritol ratio consistently associated with phosphorylated tau (pTau) and total tau (tTau), but not amyloid-β. This association was validated in an independent CSF metabolomics (n=687) and proteomics (n=737) cohorts. Structural equation modelling confirmed that polyol metabolites predict tau burden, with less than 3% attenuation following genetic adjustment, establishing a non-genetic, metabolically driven mechanism.

These findings define a tau-dominant, amyloid-independent metabolic axis in neurodegeneration, implicating the polyol pathway as a potentially modifiable therapeutic target.

## Introduction

Dementia is a major public health challenge, affecting nearly 60 million people worldwide, with this prevalence projected to rise to 152 million by 2050. This increase is expected to be disproportionately distributed, with greatest impact in the low- and middle-income countries, where diagnostic capacity and access to specialized care remains limited^1,2^. Clinically, dementia encompasses a heterogeneous group of neurodegenerative disorders, with overlapping symptoms and syndromes, complicating early diagnostics and risk stratification, particularly at prodromal stages^3,4^. Despite substantial advances in neuroimaging and fluid biomarkers^3,5^, the molecular mechanisms linking early neuropathological changes to clinical manifestations remain only partially understood, limiting our ability to identify individuals at risk.

Metabolomics has emerged as a powerful systems biology tool for characterizing the smallmolecule perturbations associated with landscape of neurodegeneration, capturing disease-relevant perturbations spanning both causal risk factors and pathological consequences^6,7^. Blood-based metabolomics offer a minimally invasive window into systemic metabolic health assessment and its relationship with dementia risk^7,8^. However peripheral metabolites may only partially reflect peripheral nervous system processes due to dilution and compartmentalization. In contrast, cerebrospinal fluid (CSF), which is in direct contact with the brain, more closely captures biochemical alterations occurring in the central nervous system^9^, making an especially informative matrix for metabolomic investigation. From the clinical perspective, CSF-resolved metabolomics has the potential to complement established biomarkers such as amyloid beta (Aβ42), phosphorylated tau (pTau) or total tau (tTau)^10^ thereby improving clinical staging, subtype differentiation, and prognostic modelling of cognitive decline.

Previous metabolomics studies have highlighted the polyol (sorbitol-fructose) pathway as being particularly relevant in dementia^11^. This pathway provides an alternative route for glucose metabolism and is activated under conditions of hyperglycemia, oxidative stress and mitochondrial dysfunction. Elevated brain tissue glucose associates with AD pathology severity and symptom onset^12,13^, and *post-mortem* AD brain shows glucose, sorbitol and fructose accumulation ^14^, consistent with a shift of glucose flux from glycolysis to polyol formation. Given the high prevalence of metabolic comorbidities, such as type 2 diabetes (T2D) or impaired glucose tolerance in older people, among whom dementia is also more prevalent, dysregulated polyol metabolism represents a biologically plausible link between metabolic diseases and dementia. Clinically, this hints to potential implications for patient risk stratification and individualized management in clinical routine practice^15,16^.

However, the role of polyols in dementia and their relationships with established biomarker phenotypes remains poorly characterized. Most metabolomics studies in dementia have focused on either peripheral blood metabolites, which are only modestly correlated with central pathology; or on CSF lipid species, amino acids and broad untargeted metabolite panels^17^, with limited attention to polyols. Consequentially, it remains unclear whether polyol metabolites provide clinically actionable information in the context of dementia.

In this study, we pursued to main aims: (i) to explore how central and peripheral metabolic alterations relate to dementia biomarkers across clinically defined diagnostic groups; and (ii) to assess whether metabolite-based signatures provide information on clinical staging beyond established biomarkers. We performed combined metabolomic profiling in plasma and CSF from individuals spanning the cognitive spectrum from (normal cognition, MCI and dementia). We systematically characterized the metabolome in each matrix and evaluated associations between metabolites and with established dementia biomarkers and clinical outcomes.

We found increased polyols as early as MCI-AD clinical stage. Neurons, microglia and cerebrovascular cells rely on constitutively active, insulin-independent transporters. As a result, intracellular glucose levels rise passively with plasma levels concentrations. When glycolytic capacity is exceeded, excess glucose preferentially diverted into the polyol pathway, a vulnerability reflected in the CSF polyol accumulation observed here. We show for the first time that this metabolic dysregulation is detectable *in vivo* in CSF across dementias and is specifically coupled to tau rather than amyloid pathology.

## Results

### Participant characteristics (Cohort)

We performed both untargeted and targeted GC/MS metabolomics on a cohort of 167 individuals, including those with no cognitive impairment (NCI, n = 33), mild-cognitive impaired due to AD (MCI-AD, n = 21) patients, subjects with dementia due to AD (Dementia-AD, n = 81) and subjects with AD and concomitant cerebrovascular disease (”mixed dementia”, MIX, n = 32) (Figure 1). The clinical cohort characteristics are summarized in Supplementary Table S1.

**Figure 1:**
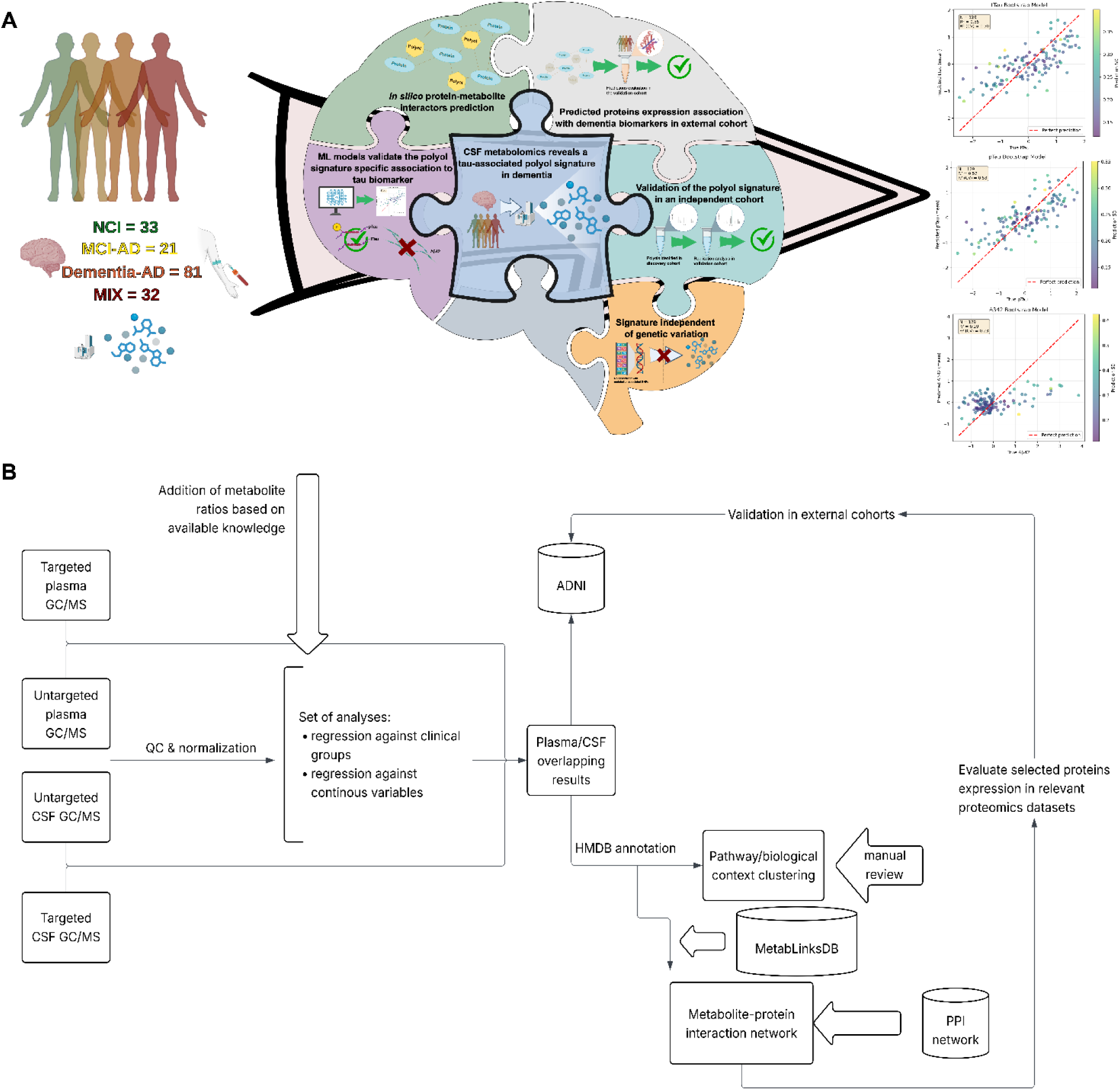
Study design, analytical framework and key outcomes of the combined CSF and plasma metabolomics study of dementia individuals. **(A)** Schematic overview of the cohort included in the study with 33 non-cognitive impaired individuals (NCI), 21 mild-cognitively impaired (MCI-AD), 81 individuals with Alzheimer’s Disease (Dementia-AD) and 21 individuals with dementia due to AD with concomitant cerebrovascular disease (MIX). A systematic metabolite-clinical biomarkers and clinical outcomes association analysis was performed, enriched with metabolite-protein interaction networks and validated in external cohorts, leading to the identification of a CSF polyol signature specifically associated with tau biomarkers. Further assessment of genetics impact upon the relevant polyol signature abundance was investigated. **(B)** Bioinformatics workflow of the metabolomics data processing and analysis, including quality assessment, normalization and association to clinical outcomes in the different analysed matrices. Further functional characterization of the relevant metabolites was performed combining HMDB annotations and metabolite-protein interaction networks. Validation of both metabolomics results and *in silico* proteins was done with datasets retrieved from the ADNI cohort.

No significant differences were found for the sex distribution across the clinical groups (p=0.235). We observed relevant differences for BMI (p=0.023), which sequentially decreased from the controls to MCI-AD and Dementia-AD while remaining higher in MIX group, as well as for age, which increased with disease severity. As expected, we observed lower MMSE score by increasing disease severity (p<0.001). The same decreasing pattern was observed for the Aβ42 protein levels, while for both pTau and tTau, the levels increased sequentially from NCI to MC-ADI and to Dementia-AD. In the MIX patients pTau and tTau levels were slightly lower than those observed in Dementia-AD patients (Figure 2A-2B). Notably, the combination of Aβ42 and tTau scores allowed for the discrimination of NCI individuals from the combined MCI-AD, Dementia-AD and MIX patients (Figure 2C).

**Figure 2:**
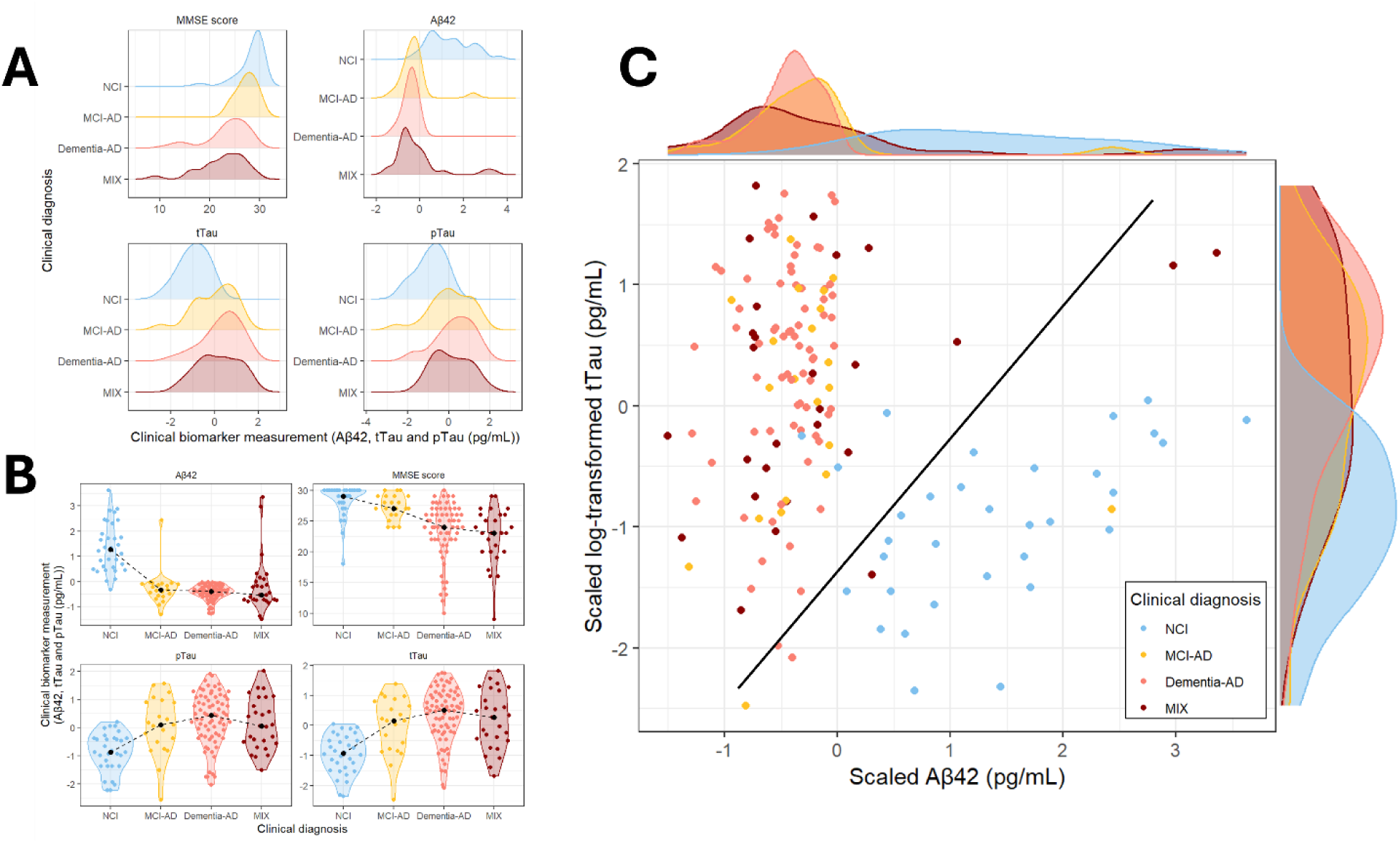
Dementia diagnostic biomarkers distribution in the cohort. (**A**) MMSE score, Aβ42 (scaled), pTau and tTau (log-transformed and scaled) scores distribution according to the dementia clinical diagnosis category. For the density distributions, plots have been coloured accordingly to the clinical diagnosis (blue = NCI; yellow = MCI-AD; pink = Dementia-AD; and red = MIX). (**B)** For the violin plots, the median value is shown as a black dot per each clinical group, with a dotted line displaying the trend. Violin plots have been coloured accordingly to the clinical group category (blue = NCI; yellow = MCI-AD; pink = Dementia-AD; and red = MIX). (**C**) Relation plot between the log-transformed Aβ42 and tTau biomarkers. Each point represents an individual, coloured accordingly to their clinical diagnosis (blue = NCI; yellow = MCI-AD; pink = Dementia-AD; and red = MIX). A diagonal line has been added to mark the differentiation between NCI individuals and dementia patients.

### Metabolomics

Untargeted plasma metabolomics detected 669 Mass Spectrometry (MS) peaks, of which 154 were annotated to known metabolites (23.02%), while the targeted metabolomics included 36 metabolites. In untargeted CSF metabolomics, 837 peaks of which were identified, with 116 (13.86%) were successfully annotated, while the targeted added 29 metabolites. After quality filtering and imputation, the final annotated dataset comprised 110 plasma and 79 CSF metabolites, supplemented by literature-informed metabolite ratios (see *Materials and Methods*).

#### Metabolites associated with neuropathology biomarkers

Across the three dementia biomarkers (Aβ42, pTau, tTau), a total of 95 metabolites showed significant associations (P<0.05), with 58 identified in CSF and 37 in plasma (Table 1).

**Table 1:** Metabolomics – dementia biomarkers summary associations. The table below summarizes the number of metabolites that were found associated to each dementia biomarker in the two analysed matrices, CSF and plasma, as well as how many of these metabolites were associated to only one biomarker (category single-association) or more than one (category multiple-association).

|  | Category | n metabolites |
| --- | --- | --- |
| Dementia biomarker | Single-association | 45 |
|  | Multiple-association | 48 |
| Aβ42 | CSF | 16 |
|  | Plasma | 21 |
| pTau | CSF | 22 |
|  | Plasma | 10 |
| tTau | CSF | 20 |
|  | Plasma | 6 |

The plasma metabolites associated with CSF-measured Aβ42 included branched-chain amino acids (BCAAs: isoleucine, leucine, valine), aromatic amino acids (phenylalanine) and gut-derived and oxidative stress-related metabolites (Extended Data Figure 1). These associations showed modest correlation scores (-0.25 – 0.23), as expected for different matrices measurements (Supplementary Table S2). In CSF (Supplementary Table S3), Aβ42 associated primarily with small carbohydrates, amino alcohols and organic acids. CSF cholesterol showed significant association with both tTau (r^2^=0.16, p-value <0.001) and pTau (r^2^=0.29, p-value <0.001), and these associations were observed in both statins’ users and non-users (Supplementary Table S4). Of particular interest, the strongest overall associations, specific to pTau and tTau, but not Aβ42, were observed for a cluster of polyol and sugar acid metabolites (meso-erythritol, D-threitol, L-arabitol, ribonic acid, 3-deoxy-erythro-pentitol, D-glucose/erythritol, sorbitol).

#### Disease categories

Across all clinical comparisons, 123 metabolites were associated with at least one diagnostic group or biomarker, including 66 in CSF, and 57 in plasma (Supplementary Table S5). In plasma, MCI-AD and Dementia-AD were both characterized by a depletion of BCAAs and their related catabolites, while MIX group showed a broader upregulation of carbohydrates, organics acids and microbial metabolites (Supplementary Table S6). In CSF, beyond a consistent reduction in BCAAs across early and late-stage dementia, the most prominent and reproducible finding was a systemic enrichment of polyol metabolites across all three clinical groups (MIC, AD and MIX) with distinct polyol species elevated at each stage (Supplementary Table S7).

#### Polyol metabolites

Taken together, the metabolomics analyses converged on polyol metabolites as the most consistently and specifically altered metabolite class in CSF across both diagnostic group comparisons and biomarker associations (Figure 3). The polyol pathway substrates ribofuranose; xylose and the ratios of d-Glucose/sorbitol and d-Glucose erythritol were differentially abundant across both the clinical groups and tau biomarkers. The polyol metabolites themselves, including 3-deoxy-erythro-pentitol; meso-erythritol, D-threitol, L-(-)-arabitol and the sorbitol related metabolites were enriched in CSF across clinical stages and showed specifically associated with tau protein. Ribonic acid, a downstream product of the polyol pathway, was also identified as relevant. Critically, these associations were specific to tau (tTau and pTau) and were not seen for Aβ42, pointing to a tau-specific neuropathological metabolic signature.

**Figure 3:**
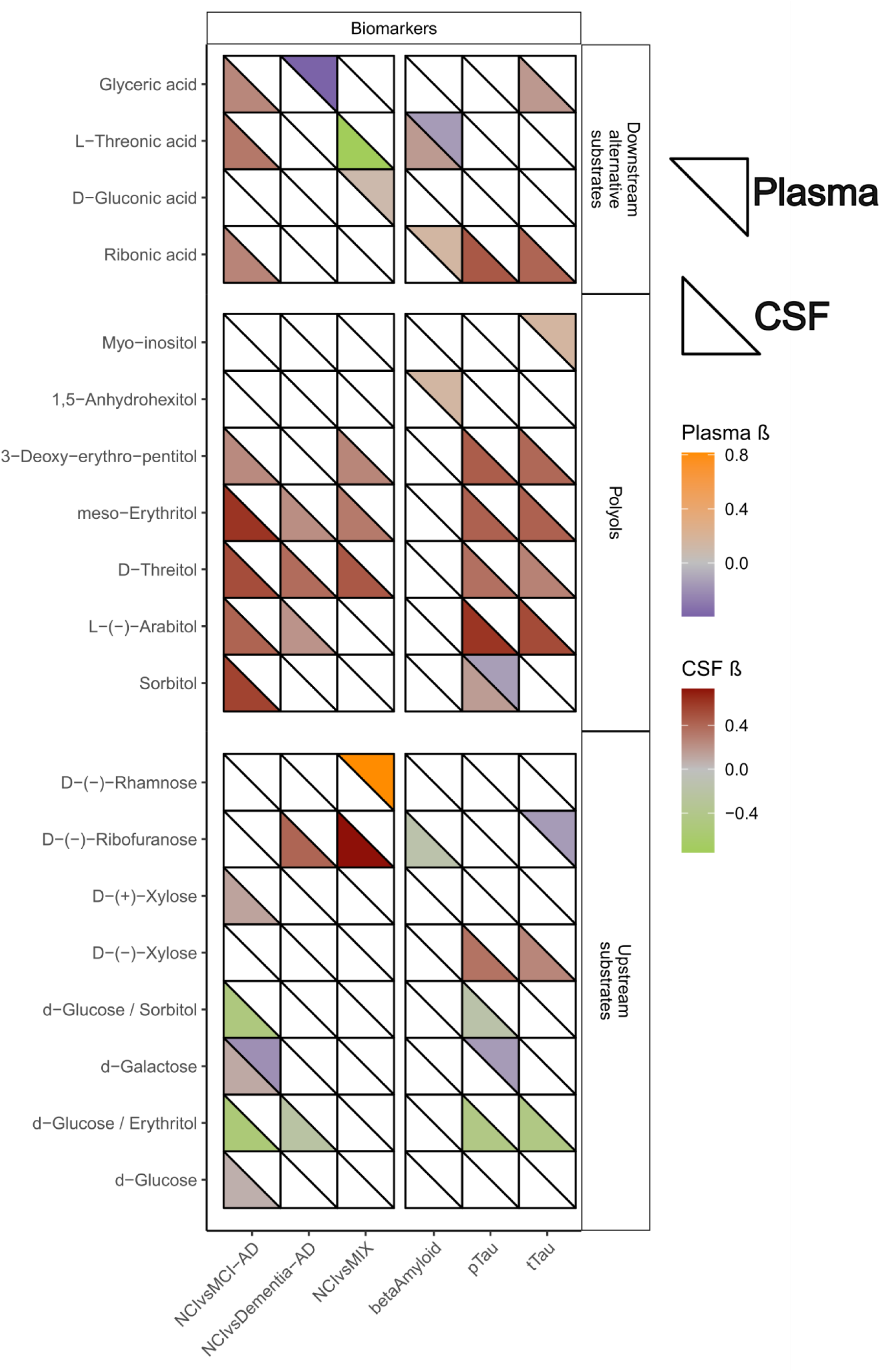
Associations between plasma and CSF polyol-related metabolites and the clinical dementia categories and common biomarkers. Polyol-related metabolite associations to tissues are represented in triangles, with the upper triangles in the tiles representing the plasma-associated metabolites and the lower triangles the CSF-associated metabolites. Associations to clinical categories are indicated with the mean beta coefficient from Bayesian models, while associations to clinical biomarkers are indicated with the beta coefficient from GLM models. Only associations that were found to be significative (p-value < 0.05) were included in the figure. The heatmap has been split in blocks according to a major classification of metabolites, from top to bottom: downstream products and/or alternative substrates; polyol metabolites themselves; and upstream substrates.

### Metabolite-protein interactions

Using HMDB-annotated metabolites and MetaLinksDB, we constructed a full metabolite-protein interaction network including all metabolites overlapping with MetaLinksDB database (n=19 metabolites) and the corresponding 461 protein interactors. In addition, a polyol-centered network was generated, which included 5 polyols (D-ribose, L-arabitol, sorbitol, D-galactose and D-glucose) mapped to 93 unique proteins (Supplementary Table S8). When restricted to CSF polyols associated with tTau, pTau or Aβ42, the subnetwork narrowed to three metabolites: D-(-)-ribofuranose, with 7 protein interactors, L-arabitol with 6 interactors and sorbitol, with19 interactions.

The network captured the core polyol pathway enzymes including aldose reductase (AKR1B1), sorbitol dehydrogenase (SORD) and AKR1A1 and AKR7A2, alongside glucose transporters (SLC2A and SLC5A) and hexokinases (HK). The involvement of hexokinases suggesting a potential suppression of glycolytic entry consistent with a metabolic shift towards polyol metabolism. Metabolite-based overrepresentation analysis identified two significantly enriched KEGG pathways: galactose metabolism (fold-enrichment = 31, adj. p-value = 0.0002) and starch and sucrose metabolism (fold-enrichment = 12.2, adj. p-value = 0.021). When expanding the analysis to include protein interactors, 41 differentially enriched pathways were identified, with galactose metabolism ranking first (fold-enrichment = 107.31, adj. p-value = 3.07x10^-41^) and, noteworthy, fructose and mannose metabolism pathway, containing the key polyol metabolite sorbitol, was among the top 10 enriched pathways (fold-enrichment = 31.22, adj. p-value = 1.33x10^-8^, Supplementary Table S9). GO:Biological Processes enrichment further identified three polyol-specific pathways: polyol transmembrane transport (fold-enrichment = 37.16, adj. p-value = 0.008), polyol catabolic process (fold-enrichment = 20.44, p-value = 0.047) and polyol metabolic process (fold-enrichment = 5.68, adj. p-value = 0.016) (Supplementary Table S9).

#### Proteins evaluation with external datasets

To validate the metabolite-protein interactors, we evaluated their expression in the Cruchaga CSF SomaScan proteomics dataset^18^ (ADNI cohort^19^), using linear regression models corrected for age and sex. Associations with tTau and pTau were substantially stronger and more consistent than those observed for Aβ42 (FDR < 0.05, Supplementary Table S10). The core polyol pathway entry enzymes AKR1B1 and AKR1B10 were positively associated with both tau biomarkers, while SORD showed a weaker positive association, coherent with the network-predicted increased polyol flux with impaired clearance (Figure 4). Hexokinases were negatively associated with both tTau and pTau, supporting the inferred diversion of glucose flux away from glycolysis, with only HK1 showing a significant association with Aβ42. Proteins involved in pentose, glycation and carbonyl stress pathways (ALDH3A1, RBKS, GALK1, NEU1, CTSA) again associated strongly with tau but not with Aβ42. None of the above associations were observed for Aβ42, reinforcing on the hypothesis of a tau-specific dysregulation of polyol metabolism.

**Figure 4:**
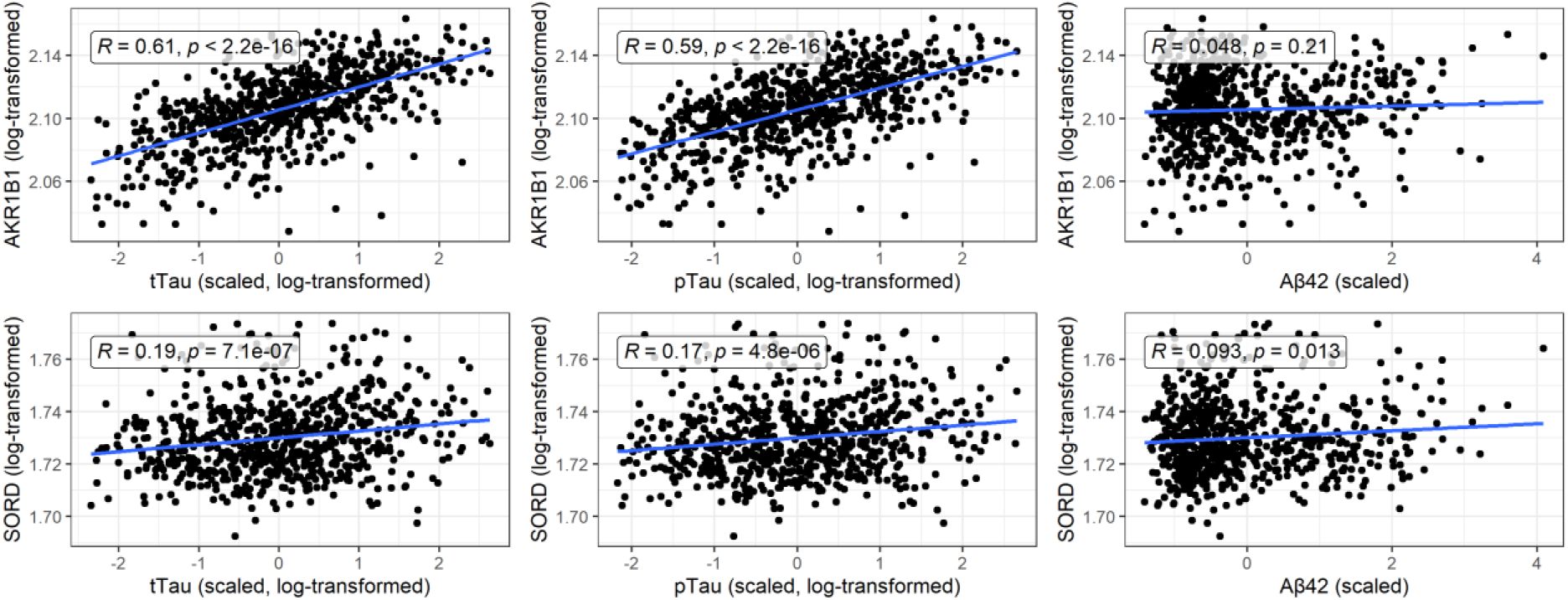
Cruchaga CSF proteomics validation. Scatter plots for the associations between the canonical polyol pathway enzymes and the CSF dementia biomarkers levels in the Cruchaga CSF SomaScan dataset. Each point represents an individual in the dataset, with the blue lines showing the fitted linear regression models.

Network coverage analyses confirmed this specificity: both tTau- and pTau-associated proteins covered over 85% of the polyol network (31/35 and 30/35 proteins, respectively; Fisher’s exact test, p = 1.5x10^-79^ and 1.9x10^-76^, respectively), whereas Aβ42-associated proteins covered only 57% of the network (20/35 proteins, p = 1.3x10^-48^).

### Metabolomics validation

We used the same Cruchaga CSF dataset^20^, for which metabolomics data was also collected, to compare our results to an external cohort. Results of these models are presented in Table 2.

**Table 2:**
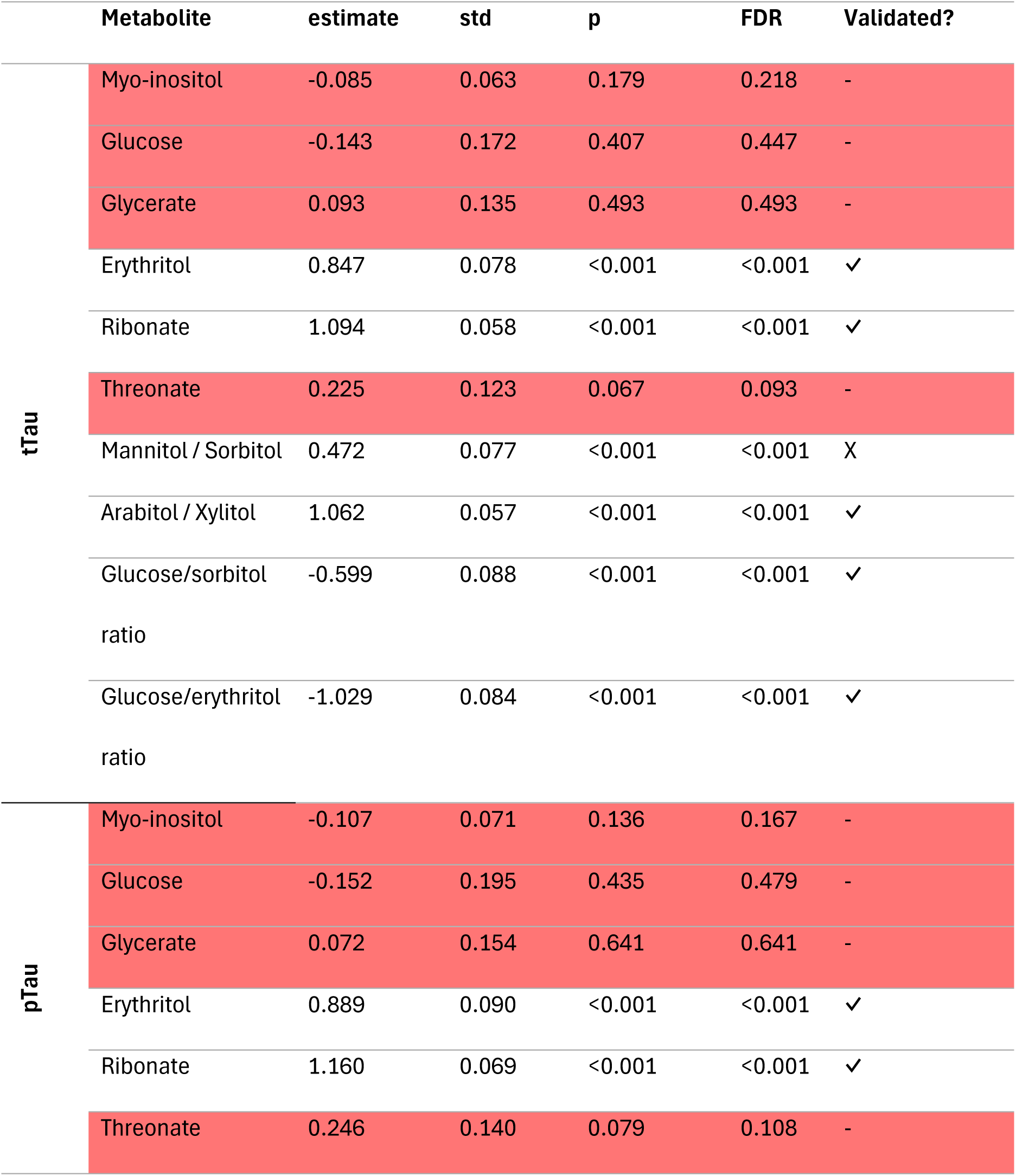
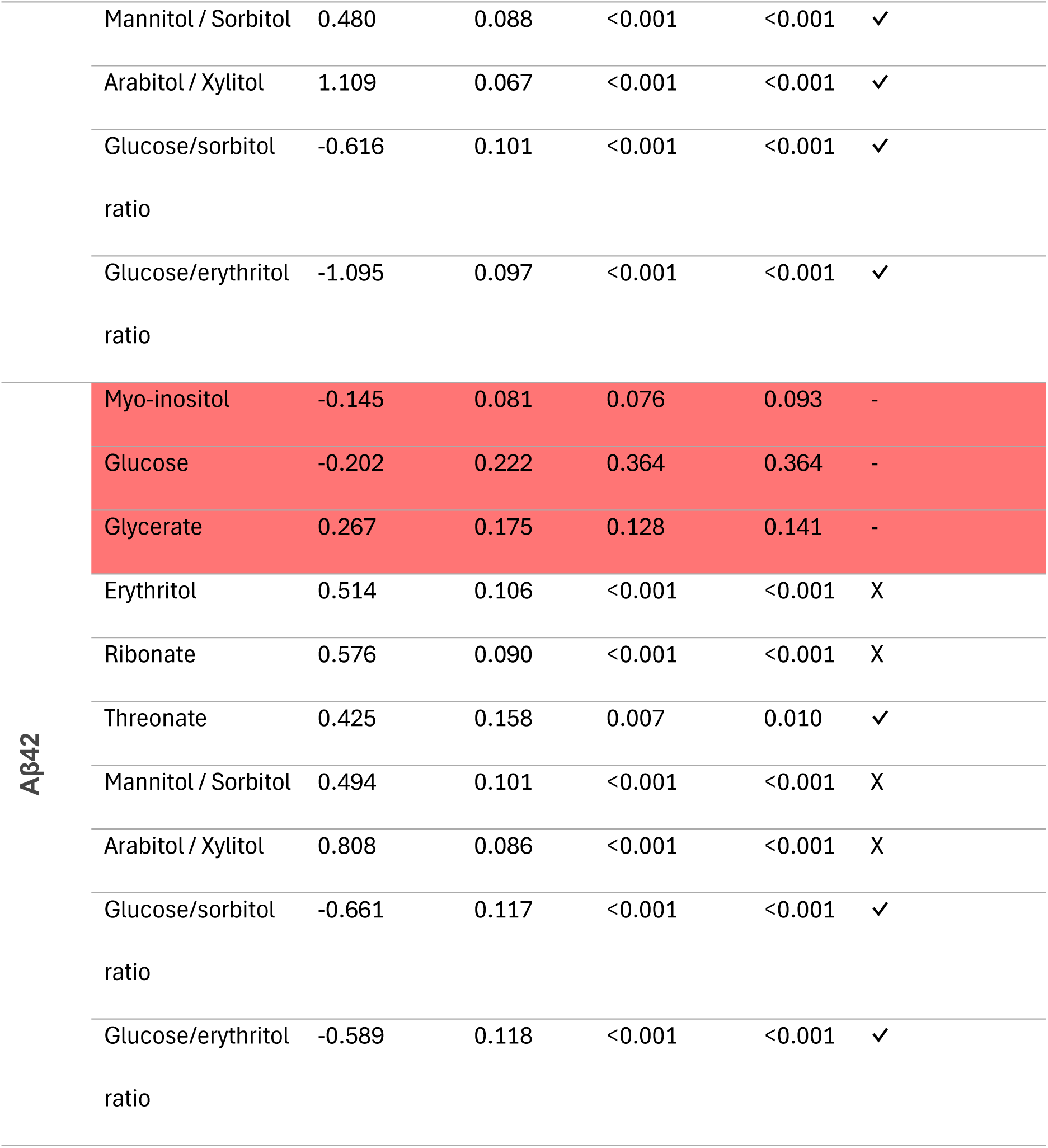
CSF polyols association to dementia biomarkers validation in Cruchaga CSF cohort. Metabolites are named as they were identified in the Cruchaga cohort. Validated column indicates whether the identified metabolite is (i) significantly associated with a dementia biomarker in Cruchaga’s cohort; and (ii) the estimate has the same directionality in both ours and Cruchaga’s cohort. For validated column, we include three symbols: “-“ if FDR > 0.05, “✓” if the metabolite complies with both estimate and FDR requirements and “X” if it failed in any of the estimates or FDR requirement.

We replicated the associations between CSF metabolites and both tTau and pTau proteins in our discovery cohort, despite the differences in methodological and analytical approaches. Ribonic acid, thionic acid, erythritol, sorbitol and xylose showed, in both cohorts, a positive association with both tTau and pTau proteins, indicating increased abundance with tau accumulation. Furthermore, glucose ratios, indicating transformation into erythritol and sorbitol were found to be negatively associated to both tTau and pTau biomarkers in both discovery and validation cohorts. In contrast, the validation of Aβ42 metabolites, instead, showed only partial replication, mostly at pathway level. These partial discrepancies were found, for example, for ribonic acid or xylose, which showed distinct association directionality in the two cohorts.

### Polyols signature model construction

To formally quantify the predictive relationship between the CSF polyol signature and AD pathology biomarkers, we built Elastic Net regression models with internal cross-validation and bootstrapped stability assesment (1,000 iterations). Consistent with all preceding analyses, the polyol signature predicted the tTau and pTau levels substantially better than Aβ42 (Figure 5A). The tTau model showed moderate predictive performance (R^2^ = 0.55, cross-validated R^2^ = 0.50) and retained eight metabolites, with L-(-)-arabitol (β=0.374), ribonic acid (β=0.123) and D-threitol (β=0.122) as the strongest contributors (Figure 5B, Supplementary Table S11). The pTau model performed similarly (R^2^=0.57, cross-validated R^2^=0.53), with an overlapping metabolite set led by L-(-)-arabitol (β=0.439), ribonic acid (β=0.167) and D-threitol (β=0.159), confirming a consitent polyol signal across both tau biomarkers (Figure 4B, Supplementary Table S11). Both models were significant under permutation testing (p-value=0.001). In contrast, the polyol signature showed poor predictive performance for Aβ42 (R^2^ =0.29, crossvalidated= 0.23), with age rather than polyol metabolites driving the model (β=-0.261). External validation in the Cruchaga CSF metabolomics cohort, despite only partial metabolite overlap, replicated moderate predictive performance for tTau (R^2^ 0.42) and pTau (R^2^ 0.36), with the same core polyols identified as top contributors (Extended Data Figure 2, Supplementary Table S12), while the Aβ42 prediction remained poor (R^2^ 0.17).

**Figure 5:**
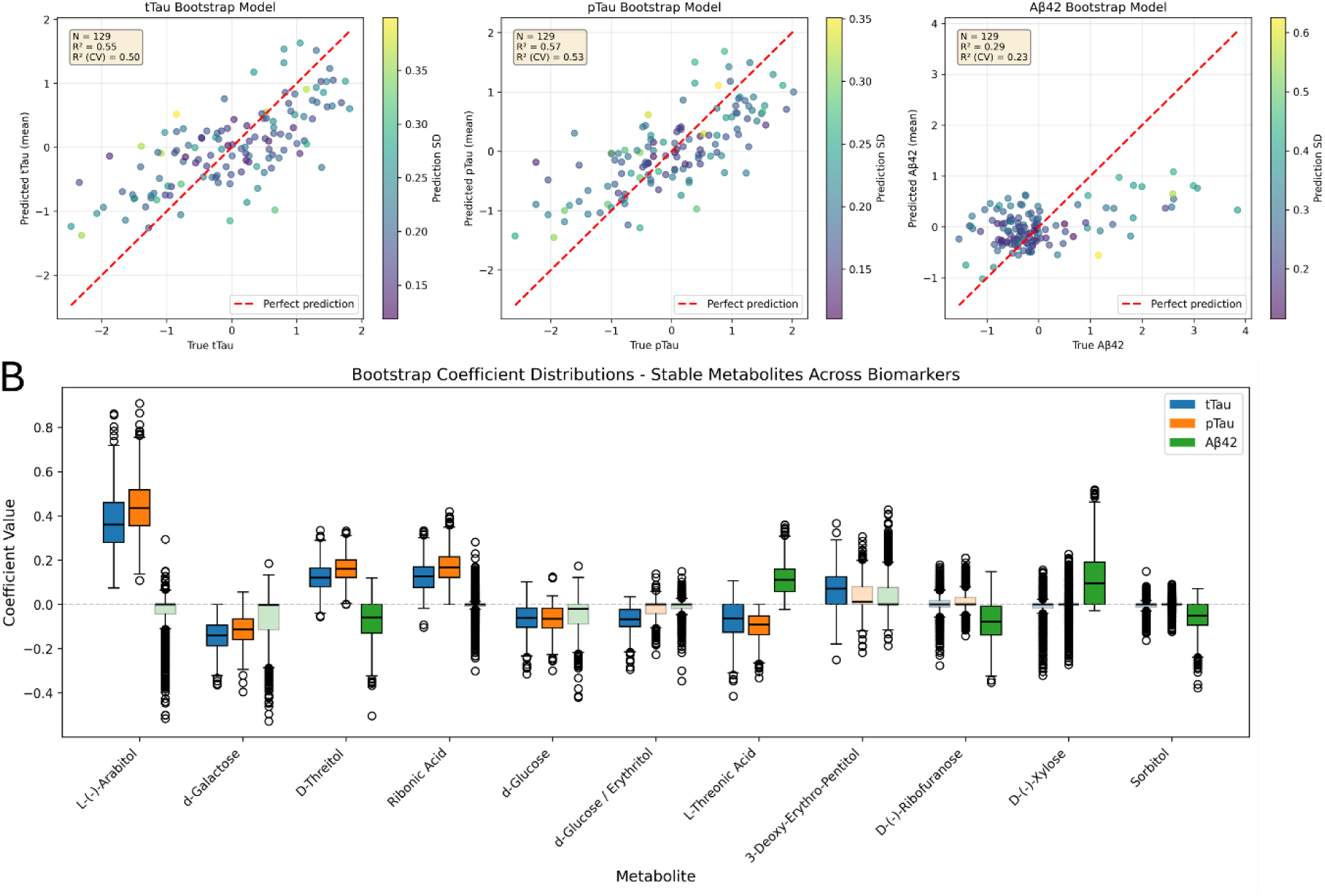
Cross-validated prediction of dementia CSF biomarkers using the polyol signature metabolite profiles. **(A)** Observed versus mean predicted values for CSF tTau, pTau, Aβ42 obtained from Elastic Net regression models adjusted for age and sex (N = 129) and 1,000 bootstrap iterations. Points represent individual participants, colored by the SD of the predictions across the 1,000 bootstrap runs, and dashed red lines indicate the identity line corresponding to perfect prediction. Model performance is summarized by the mean coefficient of determination (R²) for the full model fit and mean cross-validation (CV) as indicated in each panel. **(B)** Contributions of the tested polyol-related metabolites in the ElasticNet models for each of the three dementia biomarkers, as determined by the boxplot filling, across the bootstrap runs. Stable metabolites (>70% of selection frequency) are opaque filled, while non-stable metabolites are filled with transparency.

### Mediation evaluation and genetic adjustment

To test whether the polyol signature influences clinical diagnosis through tau pathology specifically, we applied SEM with a mediation framework. The latent polyol factor showed robust indirect effects on AD diagnosis mediated both by tTau (β=0.317, p<0.0001) and pTau (β=0.364, p<0.001), while no mediation was observed for Aβ42 (Figure 6, Supplementary Table S13), being the most contributing polyols to the models the strongest contributors identified in the ElasticNet models (Extended Data Figure 3). In MCI-AD significant total effects were observed for both tau biomarkers (tTau: β=0.301, p < 0.001; pTau: β=0.342, p = 0.004) without significant mediation, suggesting that the polyol-cognition relationship at this stage precedes or operates independently of measurable tau accumulation. These findings were replicated in the Cruchaga validation cohort for both tTau and pTau contrasts, with comparable effect sizes, while Aβ42 replication was limited by minimal metabolite overlap (Supplementary Table S14, Extended Data Figure 4).

**Figure 6:**
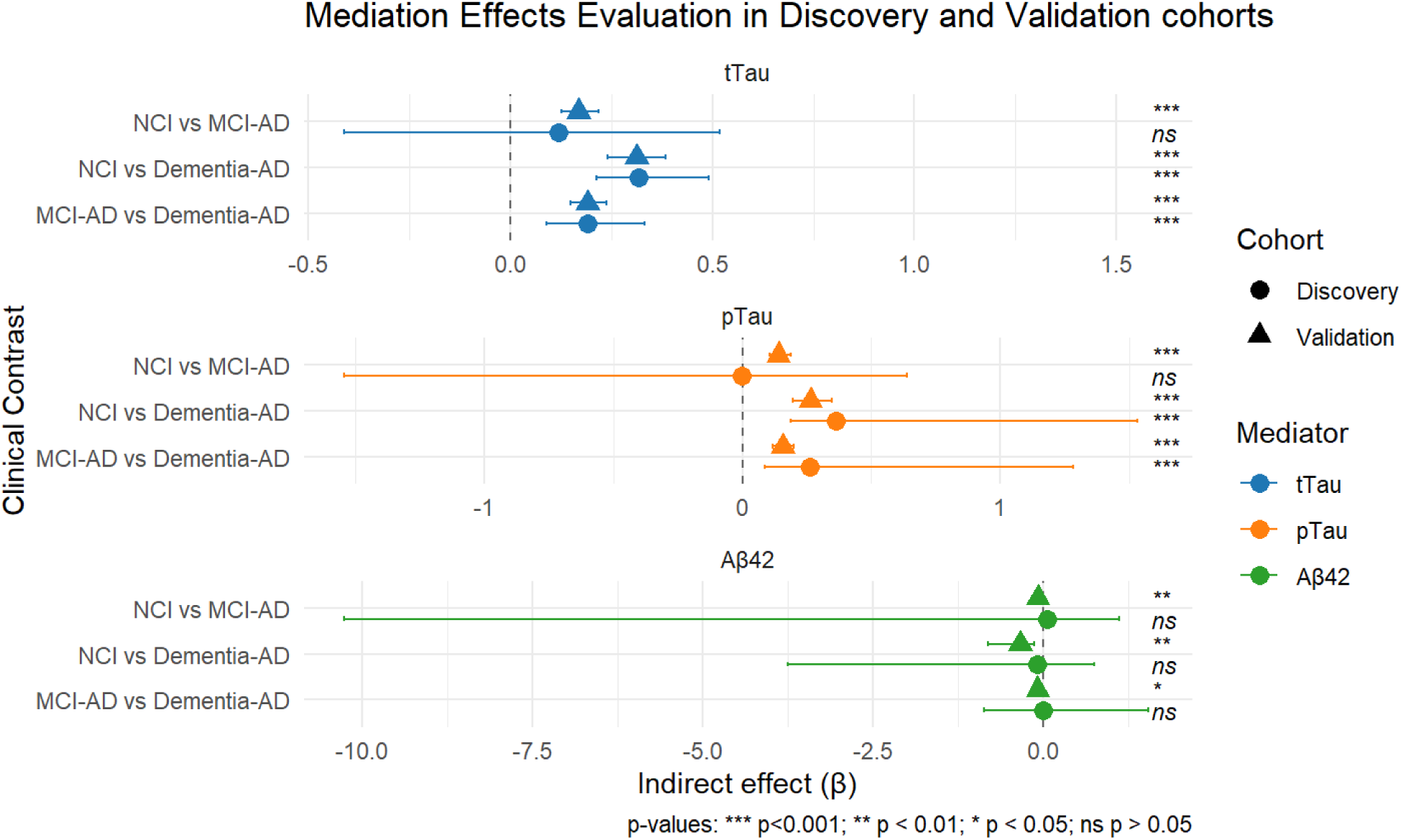
SEM results in both discovery and validation cohort for the three clinical biomarkers and contrasts. Indirect effects, presented as the median beta effect (point or triangle) and the 95% confidence intervals as horizontal bars from SEM testing polyol metabolite mediation of AD biomarker associations across clinical diagnostic groups. The discovery cohort (n=129) is presented as circles, while the validation cohort (n=) is presented in triangles for the three mediators tTau (blue), pTau (orange) and Aβ42 (green). Significance levels on the right indicate validation cohort results.

Finally, incorporation of an unweighted metabolite-linked genetic risk score (GRS, Supplementary Table S15) into the SEM models produced negligible attenuation of the indirect effects (1.0% for tTau and 2.7% for pTau), confirming that the polyol-tau-AD pathway reflects nongenetic, metabolic dysregulation rather than constitutional genetic predisposition (Figure 7, Supplementary Table S16)

**Figure 7:**
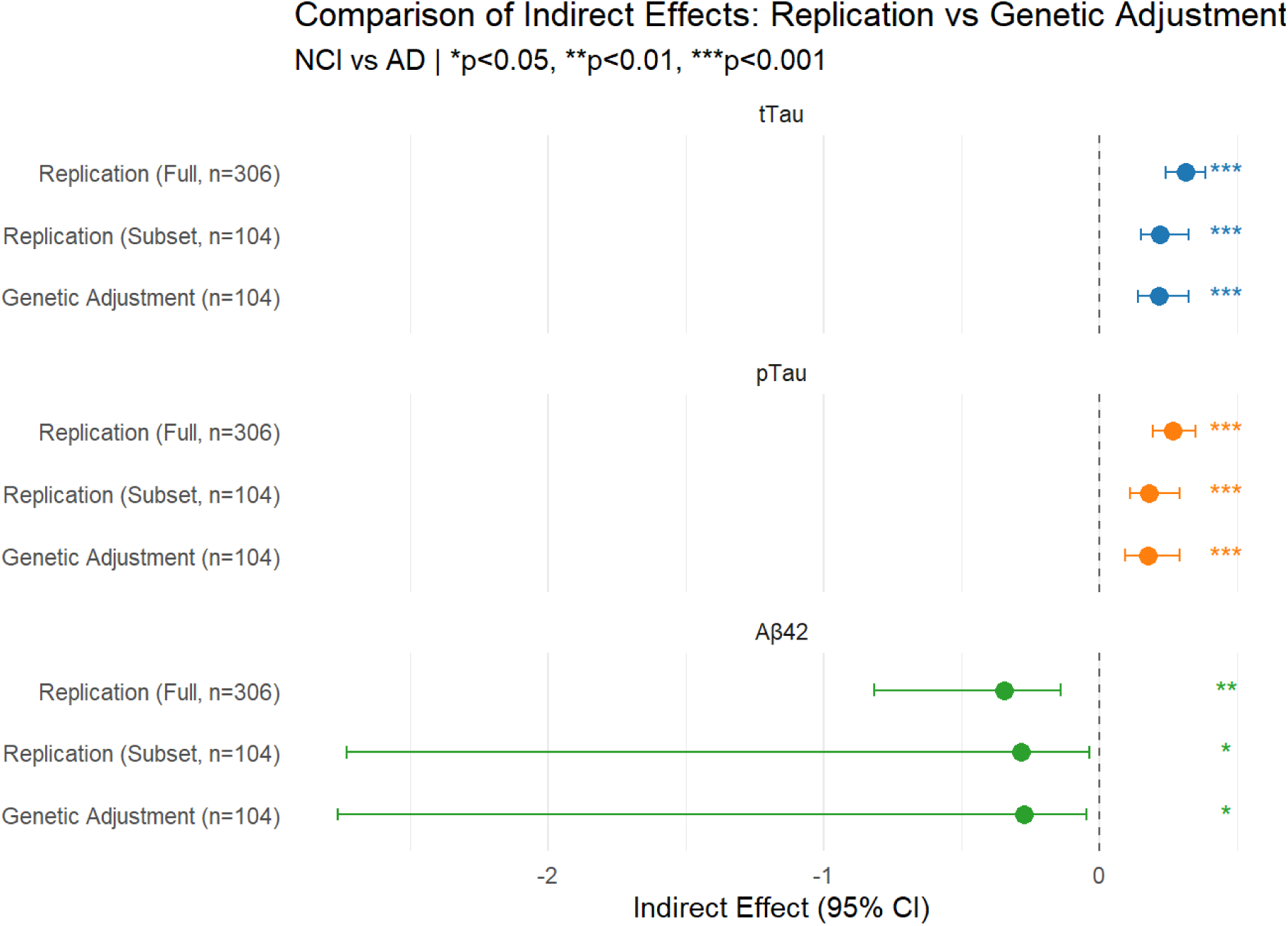
Impact of the genetics upon the polyol signature-clinical biomarkers-AD association. Comparison of indirect effects (β with 95% CI) for NCI vs Dementia-AD contrast across three analysis conditions: full validation cohort replication (n=306), genetics subset replication (n=104), and genetics subset with polyol pathway genetic risk score (GRS) adjustment (n=104). Models tested polyol metabolites → biomarker → AD mediation for tTau (blue), pTau (orange), and Aβ42 (green).

## Discussion

In this work, we report a tau-dominant metabolic axis in dementia, centered on polyol pathway dysregulation, which is independent of amyloid burden, consistent across dementia diagnoses and reproducible in external cohorts. From a clinical perspective, this is relevant for two reasons. First, the current therapeutic landscape for AD is shaped by anti-amyloid strategies. While β-amyloid monoclonal antibodies have shown some capacity to slow AD-related clinical decline^21,22^, their utility is restricted by the therapeutic window limited to early amyloid-positive individuals. Second, despite the evidence that tau burden tracks cognitive decline more faithfully than the amyloid load^8,23,24^, no tau-targeted therapy has been approved to date by any regulatory agency, although some candidates exist in both Phases 2 and 3^25^. Our data propose a metabolically tractable, pathway through which tau pathology may be driven, potentially nongenetic in origin and, therefore, potentially modifiable through pharmacological and lifestyle interventions.

The most robust and clinically interpretable findings of the study concern the central energy metabolism cluster, specifically the polyol pathway, which is seen dysregulated even in prodromal dementia (MCI-AD), coherently with the available literature^13^. The combination of elevated polyol-related metabolites (Figure 2) in CSF across multiple diagnoses, coupled to their association to both tTau and pTau biomarkers, but not Aβ42, points to a biochemically coherent and tau-specific metabolic signature^26^. These polyol metabolites and osmolytes, including myo-inositol, have previously been implicated in brain fructose metabolism and dementia-associated brain accumulation, through downstream effects on redox stress and microglial inflammation^14,27^. Strikingly, this dysregulation was detectable across all clinical stages, including MCI-AD, a patient group for which we found d-Glucose levels elevated, indicating that polyol pathway activation accompanies the earliest clinically recognizable cognitive changes, even when tau accumulation has not reached dementia levels. Clinically, this temporal position directly relates to the concept of a therapeutic window: if polyol-driven glycation stress contributes causally to tau pathology, then interventions at early stages when tau accumulation hasn’t reached the dementia defining threshold, could potentially delay or attenuate progression before irreversible tau-driven neuronal loss.

The classical polyol pathway reduces glucose to sorbitol via AKR1B1 using NADPH, and then oxidizes sorbitol to fructose through SORD using NAD^+27^. Fructose is transported into microglia, which is the principal brain-resident immune cell and a key driver of neuroinflammation in AD via the GLUT5 transporter^27,28^, where is then phosphorylated by ketohexokinase (KHK), consuming ATP^28^. This metabolic cascade drives oxidative stress through two converging mechanisms: ARK1B1 consumes NADPH, impairing glutathione recycling and amplifying reactive oxygen species (ROS) generation^29^; and KHK activity depletes ATP, generating uric acid and further ROS, ultimately triggering mitochondrial dysfunction and NF-κB–mediated neuroinflammation^27^. Additionally, polyol metabolites act as precursors for Advanced Glycation End products (AGEs), which activate RAGE receptors to drive MAPK/JNK and p38 signaling and downstream tau hyperphosphorylation^30^. The reported accumulation of polyol metabolites in *post-mortem* AD brains, even before symptom onset^14^, is consistent with our in vivo CSF observations, reinforces the pathogenic plausibility of this mechanism and adds to the clinical relevance of our findings. Furthermore, arabitol has been independently identified as a pTau consistent predictor in a large-scale CSF metabolomics study using elastic net regression across an independent cohort ^31^, further supporting the polyol signal we report.

These findings acquire clinical importance when we evaluate them through the diabetes-dementia lens and drug repurposing opportunities. While T2D confers a 59% increased risk of all-cause dementia^32^, the mechanistic explanation for this increased risk remains elusive. Our data now offers a new candidate to help clarify this link: chronic hyperglycemia in T2D is known to upregulate polyol flux through AKR1B1^33^, a key enzyme which our multi-cohort results place in the center of tau-associated neurodegeneration in the general population, not only in metabolically compromised individuals. Notably, polyol dysregulation is not unique to neurodegeneration among hyperglycaemia-related complications. As we have previously shown, serum polyols associate with future renal impairment in type 1 diabetes^34^. The recurrence of the same metabolite class across two distinct diabetic end-organ complications (kidney disease and, now, tau-driven neurodegeneration) suggests that AKR1B1-mediated polyol flux represents a shared effector pathway downstream of chronic hyperglycaemia, rather than a disease-specific finding. In fact, our prioritized proteins list analysis derived from the *in-silico* metabolite-protein interaction network generation reported positive associations of AKR1B1 with tau biomarkers. This mechanistic convergence raises the possibility that existent AKR1B1 inhibitors could be repurposed to attenuate polyol-driven pathology. Eparlestat is a potent, orally bioavailable, brain-penetrant inhibitor for both AKR1B1 and AKR1B10 approved for use in Japan for diabetic peripheral neuropathy^35^, and represents an immediately accessible candidate for clinical investigation. Its established tolerability profile and CNS penetrance makes it more suitable for brain-targeting applications than novel compounds requiring *de novo* investigation and development. Whether epalrestat can modulate CSF polyol metabolites and, ultimately, tau biomarker trajectories in at-risk individuals constitutes a testable hypothesis that our results now provide mechanistic rationale to pursue. Preliminary preclinical support exists for this proposition, as epalrestat has been shown to reduce tau protein levels and attenuate neuroinflammatory markers in a rat model of diabetic cognitive impairment^36^ and to improve cognitive performance in diabetic rats with scopolamine-induced amnesia^37^.

The CSF cholesterol levels showed robust positive associations with both tTau and pTau, consistent with reports linking cholesterol accumulation to tau conformational changes, impaired membrane curvature and promoted tau fibrillation^31,38–40^. Noteworthy, these associations were strengthened among individuals taking statins. This fact suggests a causal relationship between the cholesterol metabolism and tau-driven neurodegeneration rather than a confounded one. Our statin data suggest that follow-up work evaluating lipid-lowering strategies, specifically on tau-driven biomarker trajectories, would be valuable.

The metabolite–protein network derived from statistical associations does not, in isolation, establish molecular causality. However, integration with an independent CSF proteomics cohort provides convergent functional evidence for a coherent remodeling of sugar metabolism specifically coupled to tau pathology. Critically, proteomics abundance showed both AKR1B1 and AKR1B10, the enzymes mediating glucose entry into the polyol pathway, to be positively associated with tau biomarkers, while SORD, which facilitates polyol clearance, showed a weaker positive association, together suggesting a pattern consistent with net sorbitol accumulation. This inferred enzymatic directionality aligns with our CSF metabolomics findings and is consistent with active pathway-level engagement rather than coincidental metabolite covariation. Concomitant negative associations between canonical hexokinase and tau biomarkers are consistent with glycolytic entry suppression and redirection of glucose flux toward the polyol pathway. The observed downregulation in detoxifying enzymes (ALDH3A1, NEU1, CTSA) further support a picture of impaired carbonyl detoxification in tau-positive CSF^41^. Importantly, network saturation analyses in the Cruchaga validation dataset demonstrated strong alignment with both tTau and pTau, but only weakly with Aβ42, such that tau pathology recruited nearly the entire polyol pathway network compared with amyloid’s engagement of only a subset. This is consistent with polyol dysregulation representing downstream metabolic reprogramming driven by tau, rather than a shared upstream amyloid effect. Our reported tau-dominant pattern is independently supported by large-scale CSF proteomics and metabolomics in AD which consistently identify protein and metabolite modules more strongly associated with tau biomarkers than with amyloid ones^31,42–44^.

We validated CSF findings in an independent patient group, which confirmed that polyol metabolites were specifically linked to tau protein burden. Although not every individual metabolite replicated exactly, the overall biological pathway showed consistent results across both groups. Using a machine learning approach (Elastic Net regression), we found that polyol metabolite patterns were much better at predicting tau levels than amyloid plaques. The fact that polyols had almost no relationship with amyloid burden suggests that amyloid accumulation and polyol metabolism represent distinct biological processes. Structural equation modelling (SEM) then showed how these relationships could work mechanistically. The connection between disturbed polyol metabolism and an AD diagnosis operated primarily through tau pathology rather than directly. In other words, polyol metabolites appear to drive the metabolic conditions that fuel tau-related neurodegeneration, rather than independently causing cognitive decline. Noteworthy was the finding in patients with MCI-AD, as altered polyol metabolism was already associated with cognitive changes, further supporting our hypothesis of an early dementia therapeutic window.

The effect sizes minimal attenuation after adjustment for genetic liability provides direct evidence that the identified polyol-tau-AD mediation pathway reflects primarily non-genetic metabolic dysregulation rather than constitutional genetic predisposition. This implies that the polyol metabolic signature is, in principle, modifiable. The metabolome is the integrated readout of diet, lifestyle, environmental exposures and system metabolic health status^8^. Our findings are therefore compatible with a model in which dietary patterns promoting glycaemic dysregulation could amplify polyol pathway flux, accelerating tau pathology and downstream cognitive decline.

The principal strengths of this work are the multi-level convergence of evidence, which includes CSF metabolomics, external metabolomics validation and integration with proteomics through metabolite-protein interaction networks, as well as the consistency of the polyol signal across independent datasets and analytical approaches. The demonstration that the polyol-tau association is non-genetic, reproducible across cohorts and functionally ordered at the enzymatic level substantially elevates confidence in its biological relevance. Limitations include the crosssectional design, which precludes inference about the temporal ordering of the polyol pathway activation relative to tau deposition. Additionally, the requirement for HMDB annotation and known protein interactors constrained the scope of the networks validation and the metabolites available for cross-cohort comparison were restricted to those quantified on common platforms. Future longitudinal studies, ideally incorporating serial CSF and plasma sampling alongside dietary and glycemic phenotyping will be essential to establish whether polyol metabolite trajectories predict tau biomarker progression and, ultimately, cognitive decline at the individual patient level. Targeted pharmacological studies of AKR1B1 inhibition, such as could be epalrestat repurposing trials, represent a logical translational next step.

In summary, this study describes a tau-dominant metabolic axis in neurodegeneration, independent of amyloid burden, characterized by CSF polyol metabolites and validated through proteomics-based interaction networks across independent cohorts. The non-genetic, metabolically tractable nature of this signature, its emergency at early (MCI-AD) clinical stages, its mechanistic coherence with the diabetes-dementia axis and the existence of approved inhibitors of its key enzymatic drivers collectively position the polyol pathway as a potential clinically actionable target for tau-centered therapeutic development in dementia.

## Materials & Methods

### Discovery Phase Study

For this study, we included a subset of the participants described previously^45^. Specifically, 167 individuals were recruited from the Danish Dementia Research Centre at Copenhagen University Hospital, Rigshospitalet, following written informed consent and in accordance with the Declaration of Helsinki. Diagnosis was performed after comprehensive assessment as described in^46^, with individuals being characterized in four groups: no cognitive impairment (NCI, n=33); mild cognitive impairment (MCI-AD, n=21); Alzheimer’s disease patients (Dementia-AD, n=81); and dementia due to AD with concomitant cerebrovascular disease (MIX, n=32). Routine CSF clinical biomarkers (Aβ42, pTau and tTau) were measured for all participants as part of the diagnostics process. Plasma and CSF samples were stored at -80°C until processed. The participants were divided into Aβ positives (AD and MIX) and Aβ negatives (NCI) with a CSF cut-off for p-tau181/Aβ42 locally established at 0.077.

### Validation studies

Data used in the preparation of this article were obtained from the Alzheimer’s Disease Neuroimaging Initiative (ADNI)^47^ database (adni.loni.usc.edu). The ADNI was launched in 2003 as a public-private partnership, led by Principal Investigator Michael W. Weiner, MD. The primary goal of ADNI has been to test whether serial magnetic resonance imaging (MRI), positron emission tomography (PET), other biological markers, and clinical and neuropsychological assessment can be combined to measure the progression of mild cognitive impairment (MCI) and early Alzheimer’s disease (AD).

Specifically, we used the Cruchaga CSF metabolomics^20^ dataset, with n=687 individuals, to validate our individual metabolite results in an external cohort and test our machine-learning models generalization capabilities.

The Cruchaga’s proteomics^18^ dataset, with n=737 individuals, was used to validate our *in-silico* predicted protein metabolite interactors and their associations with dementia clinical biomarkers.

Finally, we leveraged the ADNI’s genetics data^48^, restricted to those individuals with a metabolomics profiling in the Cruchaga’s cohort described above (n=282) to test the genetics influence, through a set of pre-selected SNPs associated with polyol-related metabolites, on our identified tau-associated CSF metabolite signature.

### Metabolomics data acquisition

Metabolites were measured using both, a targeted and an untargeted mass spectrometry approach, the details of which have been published previously ^49^. In brief, both plasma and CSF samples underwent the same extraction procedure: addition of isotope-labelled internal standards, extraction via a slightly modified Bligh and Dyer liquid-liquid extraction, centrifugation, and solvent evaporation to dryness. Dried extracts were derivatized by sequential methoximation with methoxyamine hydrochloride (45°C, 60 min) and trimethylsilyl derivatization with MSTFA containing 1% TMCS (45°C, 60 min), then analysed by GC-QTOF-MS using an Agilent 8890 GC system coupled to an Agilent 7250 QTOF mass spectrometer (Agilent, Santa Clara, CA, USA). Targeted metabolites were fully quantified with calibration curves of pure standard.

### Metabolomics data analysis

#### Untargeted metabolomics

Untargeted metabolomics were first filtered to retain only annotated peaks. Peaks with higher abundance in blank samples than in biological samples (log_2_FC>0) were removed from subsequent analyses. For the remaining peaks, the mean abundance in blank samples was subtracted from the corresponding sample intensities, negative values set to missing. Any sample with more than 30% missing values, and metabolites missing more than 30% of observations were excluded. Remaining missing values were imputed using k-nearest neighbors (kNN) approach with *VIM* (v6.2.2) R package. Peak intensities were subsequently normalized using internal standards via cross-contribution robust multiple standard normalization (CRMN) method implemented in the *crmn (*v0.0.21) R package. Outlier samples were identified through injection standard evaluation and k-means clustering.

Metabolite ratios were defined *a priori* based on literature-reported metabolites across five mechanistic axes relevant to dementia: gut–brain neuroinflammation, brain energy metabolism, membrane and lipid integrity, systemic clearance, and oxidative/redox stress. Since all metabolite abundances were log10-transformed prior to analysis, pairwise ratios were computed as differences of log-transformed values (log[A/B] = log A − log B), equivalent to a logratio transformation, and appended to the metabolite matrix for all downstream analyses. In plasma (n = 16 metabolite ratios), key features included: the indole-3-propionic acid / indole-3-acetic acid ratio as an index of microbiome-derived neuroprotective output; hippuric acid / *p*-cresol and hippuric acid / phenol as proxies for host–microbial detoxification capacity; DHA / tetradecanoic acid as a membrane omega-3 tone index; palmitoleic acid / tetradecanoic acid as a desaturation index reflecting SCD1 activity and cardiometabolic risk; creatinine / urea as a renal clearance confounder; D-glucose / sorbitol to index polyol pathway flux; L-tryptophan / 5-hydroxyindoleacetic acid to capture serotonergic pathway flux; and α-tocopherol / β-tocopherol as an antioxidant status marker. Additional ratios captured BCAA catabolism, nitrogen waste handling, lipid chain-length balance, and carboxylic acid pathway flux.

In CSF (n = 26 metabolite ratios), ratios were designed to reflect CNS-intrinsic metabolic axes. The 3-hydroxybutyrate / pyruvic acid ratio indexed the shift from glucose to ketone body utilisation as an alternate brain fuel, a compensatory mechanism observed in early AD. Phospholipid membrane turnover was captured by O-phosphoethanolamine / phosphoric acid; CSF desaturation indices by palmitoleic acid TMS / tetradecanoic acid and myristoleic acid / tetradecanoic acid; and polyol pathway activation by D-glucose / sorbitol and D-glucose / *meso*-erythritol. Purine oxidative load relative to antioxidant capacity was indexed by uric acid / urea and uric acid / creatinine. Additional CSF ratios covered TCA cycle anaplerosis, urea cycle activity, neurotransmitter precursor cycling, and aromatic acid oxidative balance.

#### Targeted metabolomics

For the targeted metabolomics dataset, calibration curves were first evaluated and metabolites displaying unexpected calibration behavior were flagged and excluded from further analysis. Samples with more than 30% missing values and metabolite missing more than 30% of observations were removed. Remaining missing values were imputed using kNN approach using the *VIM* (v6.2.2) R package. Data was subsequently log-transformed to approximate normality. Outlier samples were identified through injection standard evaluation and k-means clustering.

### Statistical analysis and models generation

#### Clinical diagnostics and biomarker associations

For both plasma and CSF metabolomics datasets (targeted and untargeted), we tested associations with clinical diagnostic groups (NCI vs MCI-AD, NCI vs Dementia-AD, and NCI vs MIX) and dementia biomarkers (tTau, pTau and AΒ42). For clinical group comparisons, we performed metabolite-wise Bayesian differential abundance analyses using hierarchical regression models implemented in the *brms* R package (v2.22.0). Metabolite abundance was specified as the predictor and diagnostic group as the outcome, adjusting for age, sex and statin use.

Weakly informative priors were specified for regression coefficients (normal(0, 1)), the intercept (normal(0, 5)), and residual variance (Student-t(3, 0, 1)), reflecting conservative assumptions about effect sizes while allowing flexibility for metabolomic data. Models were estimated using two Markov chains with 2,000 iterations each, and posterior inference was based on the full posterior distribution of the group effect. Differential abundance was assessed using posterior probabilities, with metabolites considered credibly altered when the posterior probability of the effect being consistently positive or negative exceeded 0.95.

Associations for continuous dementia biomarkers were evaluated using linear regression models. For each metabolite, a Gaussian linear model was fitted with the standardized biomarker as the outcome and the standardized metabolite abundance as the predictor, adjusted for age, sex and statin use. Effect sizes were reported as standardized regression coefficients with corresponding standard errors and 95% confidence intervals. Statistical significance was assessed using Wald tests, and false discovery rate (FDR) correction was applied across metabolites using the Benjamini–Hochberg procedure.

Human Metabolome Database (HMDB) was used to further annotate metabolites by retrieving metabolite class, subclass and corresponding HMDB and KEGG accession identifiers through approximate name matching. Annotated metabolites were subsequently used for metabolite-based functional enrichment analysis against RaMP-DB derived ontologies filtered for KEGG pathways. Metabolite overrepresentation analysis was performed with *clusterProfiler* (v4.16.0) R package.

#### Metabolite – protein networks construction and results validation

HMDB annotated metabolites were used to retrieve their protein interactors from MetaLinksDB^50^ through the *OmnipathR* R package (v3.17.7). Protein-protein interactions were retrieved from the STRING database (version 12.0 ^51^), using the human dataset and filtering by minimal confidence score ≥ 400. Proteins included in the metabolite-protein network were used for extended functional enrichment analysis in a protein-based approach. We used *clusterProfiler* (v4.16.0) R package to perform overrepresentation analysis against Gene Ontology Biological Pathways database.

Metabolites in the prioritized network and their protein interactors were validated using the independent external cohorts described before. Specifically, we leveraged the Cruchaga CSF metabolomics^20^and proteomics^18^ datasets from the Alzheimer’s Disease Neuroimaging Initiative (ADNI). For metabolite validation, the external cohort was restricted to annotated polyols overlapping with those identified in our primary analysis. Metabolomics data was log-transformed to reduce skew prior to modelling. Linear regression models were then constructed to assess associations between normalized metabolite abundances and CSF tTau, pTau, and Aβ42 levels, adjusting for age and sex as covariates. Proteomic validation in the Cruchaga CSF cohort followed an analogous approach, using protein abundances as predictors of CSF biomarker levels while controlling for age and sex. These analyses were conducted using the *limma* R package (v3.66.0). All statistical tests were two-sided, and p-values were corrected for multiple testing using the FDR approach.

#### Machine learning models generation

Elastic Net regression models were generated using scikit-learn (v1.8.0) to model CSF pTau, tTau and Aβ42 using the pre-selected CSF polyol metabolites, with age and sex as confounding factors. Both pTau and tTau were log-transformed and all biomarkers were standardized to zero mean and unit variance. Regularization parameters were optimized via cross-validation. Model performance was assessed using cross-validated coefficients of determination (R²), root mean squared error (RMSE) and mean absolute error (MAE). Statistical significance was evaluated using permutation testing with 1,000 permutations. To ensure robustness of metabolite selection, bootstrap resampling was performed with 1,000 iterations. Stable predictors were defined as metabolites selected in more than 70% of bootstrap iterations. P-values and confidence intervals for model statistics and regression coefficients were derived from the bootstrap distributions. Standardized regression coefficients were extracted to assess relative feature contributions and to enable direct comparison of effect sizes across biomarkers. Models were validated against the same Cruchaga CSF metabolomics cohort used previously^20^ which was used as a validation dataset.

#### Structural Equation Modelling and genetics influence evaluation

Structural Equation Modelling (SEM) was performed in Python using *semopy* (v2.3.11) to test if a latent polyol factor mediated the associations between dementia biomarkers and clinical diagnostics, taken as pairwise contrasts. Age and sex were included as covariates, while tTau, pTau were log-transformed and, together with Aβ42, standardized. To ensure numerical stability and model convergence under maximum likelihood estimation, all continuous variables included in each model (metabolites, biomarker mediator, age) were Z-standardized. Mediation models simultaneously estimated paths from the latent factor, containing a set of predefined polyols, to the biomarker (a-path), from the biomarker to diagnosis (b-path), and a direct path from the latent factor to diagnosis (c′), adjusting for age and sex. Indirect (a×b), direct, and total effects were derived using 1,000 bootstrap resamples to obtain median estimates, percentile-based 95% confidence intervals, and two-sided p-values. Factor loadings were summarized across bootstrap iterations and corrected for multiple testing using Benjamini–Hochberg FDR. Latent factor scores were extracted from the fitted model and used in logistic regression models to compare discriminative performance (ROC/AUC) of biomarker-only, metabolite-only, and combined models including the biomarker and the relevant polyols.

The Cruchaga CSF metabolomics cohort was used as an external validation dataset for the SEM analyses, applying the same structural framework as defined above. Because ADNI2/GO provides genotype data on a subset of individuals, we identified single nucleotide polymorphisms (SNPs) previously reported in metabolome-wide association studies (mGWAS) to be associated with the levels of polyols of interest. These SNPs were extracted from genotyped individuals in the validation cohort and used to construct an unweighted genetic risk score (GRS), due to the lack of GWAS summary statistics for the ADNI cohort. For each study participant with genetics data available, we computed the GRS score by summing the individual SNP effect alleles in the ADNI2/GO dataset^19^. For each clinical contrast and dementia biomarker, similar metabolite panels, based on available overlapping metabolites, were used to construct the latent polyol factor within a confirmatory SEM framework. Two validation models were tested: (1) replication without genetic adjustment; and (2) genetic adjustment including an unweighted GRS as a covariate in mediator and outcome models. All continuous predictors (metabolites, biomarker, age, and GRS where applicable) were z-standardized to ensure numerical stability. Indirect, direct, and total effects were estimated using maximum likelihood and 1,000 bootstrap resamples to derive median estimates, percentile-based 95% confidence intervals, p-values, and model convergence rates. Results from the full metabolomics dataset and the genetics subset were compared to assess representativeness, and attenuation of the indirect effect after GRS adjustment was quantified to evaluate potential genetic confounding versus independent metabolic pathways.

All statistical analyses were performed in R (v4.5.0, https://www.R-project.org/) and Python (v3.13.17).

## Data Availability

The dataset analyzed here is not directly publicly available, for the privacy of the participants, in compliance with EU and Danish data protection law. The data can be accessed upon rea-sonable request and relevant legal permission from the Danish data protection agency. Data access requests should be directed to S.G.H. Validation cohort data are publicly accessible through the repositories referenced in the manuscript.
All necessary scripts to reproduce the analyses reported here are available at: github.com/ SDCC-ClinicalOmics/dementia_CSF_polyols.

## Statements

### Acknowledgments

We would like to acknowledge the participants from the Danish Dementia Center at Copenhagen University Hospital, Rigshospitalet for making this study possible.

Data collection and sharing for this project was funded by the Alzheimer’s Disease Neuroimaging Initiative (ADNI) (National Institutes of Health Grant U01 AG024904) and DOD ADNI (Department of Defense award number W81XWH-12-2-0012). Data collection and sharing for this project was funded by the Alzheimer’s Disease Metabolomics Consortium (National Institute on Aging R01AG046171, RF1AG051550 and 3U01AG024904-09S4). Data generation and sharing for this project was funded, in part, by the Alzheimer’s Disease Sequencing Project. ADNI is funded by the National Institute on Aging, the National Institute of Biomedical Imaging and Bioengineering, and through generous contributions from the following: AbbVie, Alzheimer’s Association; Alzheimer’s Drug Discovery Foundation; Araclon Biotech; BioClinica, Inc.; Biogen; Bristol-Myers Squibb Company; CereSpir, Inc.; Cogstate; Eisai Inc.; Elan Pharmaceuticals, Inc.; Eli Lilly and Company; EuroImmun; F. Hoffmann-La Roche Ltd and its affiliated company Genentech, Inc.; Fujirebio; GE Healthcare; IXICO Ltd.; Janssen Alzheimer Immunotherapy Research & Development, LLC.; Johnson & Johnson Pharmaceutical Research & Development LLC.; Lumosity; Lundbeck; Merck & Co., Inc.; Meso Scale Diagnostics, LLC.; NeuroRx Research; Neurotrack Technologies; Novartis Pharmaceuticals Corporation; Pfizer Inc.; Piramal Imaging; Servier; Takeda Pharmaceutical Company; and Transition Therapeutics. The Canadian Institutes of Health Research is providing funds to support ADNI clinical sites in Canada. Private sector contributions are facilitated by the Foundation for the National Institutes of Health (www.fnih.org). The grantee organization is the Northern California Institute for Research and Education, and the study is coordinated by the Alzheimer’s Therapeutic Research Institute at the University of Southern California. ADNI data are disseminated by the Laboratory for Neuro Imaging at the University of Southern California.

### Author contributions

Contributions to this article follows the <u>Contributor Role Taxonomy</u> (CRediT). **M.C-G**: Conceptualization, Formal analysis, Methodology, Project administration, Software, Visualization, Writing - original draft, Writing - review & editing. **A.W**.: Conceptualization, Formal analysis, Methodology, Project administration, Software, Visualization, Writing - original draft, Writing - review & editing. **T.M**: Formal analysis, Methodology, Software, Visualization, Writing - review & editing. **K.H**.: Formal analysis, Methodology, Project administration, Writing - review & edit- ing. **A.H.S**.: Data curation, Resources, Writing – review & editing. **L.W**.: Writing - review & editing**. P.P.:** Writing - review & editing. **R.E.M.:** Funding acquisition, Writing - review & editing. **T.S.A:** Supervision, Writing – original draft, Writing - review & editing. **T.K.:** Writing - review & editing. **S.G.H.:** Funding acquisition, Project administration, Data curation, Resources, Writing - review & editing. **C.L.-Q.**: Conceptualization, Funding acquisition, Project administration, Supervision, Writing – original draft, review & editing.

### Data and code availability

The dataset analyzed here is not directly publicly available, for the privacy of the participants, in compliance with EU and Danish data protection law. The data can be accessed upon reasonable request and relevant legal permission from the Danish data protection agency. Data access requests should be directed to S.G.H. Validation cohort data are publicly accessible through the repositories referenced in the manuscript.

All necessary scripts to reproduce the analyses reported here are available at: github.com/ SDCC-ClinicalOmics/dementia_CSF_polyols.

### Ethical approval

Protocols and procedures were approved by the Danish Health Research Ethics Committee B for Capital Region Denmark, with reference number: H-21051757.

## References

1. Nichols, E. et al. Estimation of the global prevalence of dementia in 2019 and forecasted prevalence in 2050: an analysis for the Global Burden of Disease Study 2019. Lancet Public Health 7, e105–e125 (2022).

2. Aarsland, D. et al. Prevalence of Alzheimer’s disease pathology in the community. Nature 10.1038/s41586-025-09841-y (2025) doi:10.1038/s41586-025-09841-y.

3. Sirkis, D. W., Bonham, L. W., Johnson, T. P., La Joie, R. & Yokoyama, J. S. Dissecting the clinical heterogeneity of early-onset Alzheimer’s disease. Molecular Psychiatry vol. 27 2674–2688 Preprint at 10.1038/s41380-022-01531-9 (2022).

4. Duara, R. & Barker, W. Heterogeneity in Alzheimer’s Disease Diagnosis and Progression Rates: Implications for Therapeutic Trials. Neurotherapeutics vol. 19 8–25 Preprint at 10.1007/s13311-022-01185-z (2022).

5. Zetterberg, H. & Bendlin, B. B. Biofluid biomarkers in Alzheimer’s disease and other neurodegenerative dementias. Nature 650, 49–59 (2026).

6. Jia, L. et al. A metabolite panel that differentiates Alzheimer’s disease from other dementia types. Alzheimer’s and Dementia 18, 1345–1356 (2022).

7. Zhang, X. et al. Plasma metabolomic profiles of dementia: a prospective study of 110,655 participants in the UK Biobank. BMC Med. 20, (2022).

8. You, J. et al. Mapping the plasma metabolome to human health and disease in 274,241 adults. Nat. Metab. 7, 2366–2384 (2025).

9. Maji, S. K., Anoop, A., Singh, P. K. & Jacob, R. S. CSF biomarkers for Alzheimer’s disease diagnosis. International Journal of Alzheimer’s Disease Preprint at 10.4061/2010/606802 (2010).

10. Berezhnoy, G., Laske, C. & Trautwein, C. Metabolomic profiling of CSF and blood serum elucidates general and sex-specific patterns for mild cognitive impairment and Alzheimer’s disease patients. Front. Aging Neurosci. 15, (2023).

11. Rosales-Corral, S., Tan, D. X., Manchester, L. & Reiter, R. J. Diabetes and alzheimer disease, two overlapping pathologies with the same background: Oxidative stress. Oxidative Medicine and Cellular Longevity vol. 2015 Preprint at 10.1155/2015/985845 (2015).

12. Benn, M., Nordestgaard, B. G., Tybjærg-Hansen, A. & Frikke-Schmidt, R. Impact of glucose on risk of dementia: Mendelian randomisation studies in 115,875 individuals. Diabetologia 63, 1151–1161 (2020).

13. An, Y. et al. Evidence for brain glucose dysregulation in Alzheimer’s disease. Alzheimer’s and Dementia 14, 318–329 (2018).

14. Xu, J. et al. Elevation of brain glucose and polyol-pathway intermediates with accompanying brain-copper deficiency in patients with Alzheimer’s disease: Metabolic basis for dementia. Sci. Rep. 6, (2016).

15. Zheng, J. et al. Evaluating the efficacy and mechanism of metformin targets on reducing Alzheimer’s disease risk in the general population: a Mendelian randomisation study. Diabetologia 65, 1664–1675 (2022).

16. Wium-Andersen, I. K., Osler, M., Jørgensen, M. B., Rungby, J. & Wium-Andersen, M. K. Antidiabetic medication and risk of dementia in patients with type 2 diabetes: A nested case-control study. Eur. J. Endocrinol. 181, 499–507 (2019).

17. Lista, S. et al. Integrative metabolomics science in Alzheimer’s disease: Relevance and future perspectives. Ageing Research Reviews vol. 89 Preprint at 10.1016/j.arr.2023.101987 (2023).

18. Ali, M. et al. Multi-cohort cerebrospinal fluid proteomics identifies robust molecular signatures across the Alzheimer disease continuum. Neuron 113, 1363–1379.e9 (2025).

19. 19. Beckett, L. A. et al. The Alzheimer’s Disease Neuroimaging Initiative phase 2: Increasing the length, breadth, and depth of our understanding. Alzheimer’s and Dementia vol. 11 823–831 Preprint at 10.1016/j.jalz.2015.05.004 (2015).

20. Wang, C. et al. Genetic architecture of cerebrospinal fluid and brain metabolite levels and the genetic colocalization of metabolites with human traits. Nat. Genet. 56, 2685 (2024).

21. C.H. van Dyck et al. Lecanemab in early Alzheimer’s Disease. N. Engl. J. Med. 388, 142–143 (2021).

22. Sims, J. R. et al. Donanemab in Early Symptomatic Alzheimer Disease: The TRAILBLAZER-ALZ 2 Randomized Clinical Trial. JAMA 330, 512–527 (2023).

23. Ossenkoppele, R. et al. Amyloid and tau PET-positive cognitively unimpaired individuals are at high risk for future cognitive decline. Nat. Med. 28, 2381–2387 (2022).

24. Karikari, T. K. et al. Blood phospho-tau in Alzheimer disease: analysis, interpretation, and clinical utility. Nature Reviews Neurology vol. 18 400–418 Preprint at 10.1038/s41582-022-00665-2 (2022).

25. Huang, L. K., Kuan, Y. C., Lin, H. W. & Hu, C. J. Clinical trials of new drugs for Alzheimer disease: a 2020–2023 update. Journal of Biomedical Science vol. 30 Preprint at 10.1186/s12929-023-00976-6 (2023).

26. Hotta, N. New concepts and insights on pathogenesis and treatment of diabetic complications: polyol pathway and its inhibition. Nagoya J. Med. Sci 60, 89–100 (1997).

27. Sturno, A. M., Hassell, J. E., Lanaspa, M. A. & Bruce, K. D. Do microglia metabolize fructose in Alzheimer’s disease? Journal of Neuroinflammation vol. 22 Preprint at 10.1186/s12974-025-03401-x (2025).

28. Srinivasan, K. et al. Alzheimer’s Patient Microglia Exhibit Enhanced Aging and Unique Transcriptional Activation. Cell Rep. 31, (2020).

29. Tang, W. H., Martin, K. A. & Hwa, J. Aldose reductase, oxidative stress, and diabetic mellitus. Front. Pharmacol. 3 MAY, (2012).

30. Olivera Santa-Catalina, M., et al. JNK signaling pathway regulates sorbitol-induced Tau proteolysis and apoptosis in SH-SY5Y cells by targeting caspase-3. Arch. Biochem. Biophys. 636, 42–49 (2017).

31. Francia, M. et al. Integrative analysis of cerebrospinal fluid biomarkers, metabolomics, and polygenic risk reveals novel metabolite associations with Alzheimer’s disease. Journal of Alzheimer’s Disease 108, 1677–1695 (2025).

32. Cao, F. et al. The relationship between diabetes and the dementia risk: a meta-analysis. Diabetology and Metabolic Syndrome vol. 16 Preprint at 10.1186/s13098-024-01346-4 (2024).

33. Syamprasad, N. P., et al. AKR1B1 drives hyperglycemia-induced metabolic reprogramming in MASLD-associated hepatocellular carcinoma. JHEP Reports 6, (2024).

34. Tofte, N. et al. Metabolomic Assessment Reveals Alteration in Polyols and Branched Chain Amino Acids Associated With Present and Future Renal Impairment in a Discovery Cohort of 637 Persons With Type 1 Diabetes. Front. Endocrinol. (Lausanne). 10, (2019).

35. Bailly, C. Moving toward a new horizon for the aldose reductase inhibitor epalrestat to treat drug-resistant cancer. European Journal of Pharmacology vol. 931 Preprint at 10.1016/j.ejphar.2022.175191 (2022).

36. Jaiswal, S., Mishra, S., Torgal, S. S. & Shengule, S. Neuroprotective effect of epalrestat mediated through oxidative stress markers, cytokines and TAU protein levels in diabetic rats. Life Sci. 207, 364–371 (2018).

37. Kulkarni, U. D., Kumari Kamalkishore, M., Vittalrao, A. M. & Kumar Siraganahalli Eshwaraiah, P. Cognition enhancing abilities of vitamin D, epalrestat and their combination in diabetic rats with and without scopolamine induced amnesia. Cogn. Neurodyn. 16, 483–495 (2022).

38. Jahn, T. et al. Cholesterol metabolites and plant sterols in cerebrospinal fluid are associated with Alzheimer’s cerebral pathology and clinical disease progression. Journal of Steroid Biochemistry and Molecular Biology 205, (2021).

39. El Mammeri, N., Gampp, O., Duan, P. & Hong, M. Membrane-induced tau amyloid fibrils. Commun. Biol. 6, (2023).

40. Shin, K. C., Ali Moussa, H. Y. & Park, Y. Cholesterol imbalance and neurotransmission defects in neurodegeneration. Experimental and Molecular Medicine vol. 56 1685–1690 Preprint at 10.1038/s12276-024-01273-4 (2024).

41. Picklo, M. J., Montine, T. J., Amarnath, V. & Neely, M. D. Carbonyl toxicology and Alzheimer’s disease. Toxicology and Applied Pharmacology vol. 184 187–197 Preprint at 10.1006/taap.2002.9506 (2002).

42. Wang, Z. et al. Cerebrospinal fluid proteomics identification of biomarkers for amyloid and tau PET stages. Cell Rep. Med. 6, (2025).

43. Wojtas, A. M. et al. Proteomic changes in the human cerebrovasculature in Alzheimer’s disease and related tauopathies linked to peripheral biomarkers in plasma and cerebrospinal fluid. Alzheimer’s and Dementia 20, 4043–4065 (2024).

44. Dong, R. et al. CSF metabolites associated with biomarkers of Alzheimer’s disease pathology. Front. Aging Neurosci. 15, (2023).

45. Muk, T. et al. Steroid Hormones in Dementia: A Cross-Diagnostic Molecular Analysis of Blood and Cerebrospinal Fluid. medRxiv 10.64898/2026.02.12.26346149 (2026) doi:10.64898/2026.02.12.26346149.

46. Hasselbalch, S. G. et al. The Danish Dementia Research Centre: Integrating patient care, clinical research, and national educational services. Journal of Alzheimer’s Disease vol. 107 883–898 Preprint at 10.1177/13872877251365195 (2025).

47. Weiner, M. W., et al. The Alzheimer’s Disease Neuroimaging Initiative: Progress report and future plans. Alzheimer’s and Dementia 6, (2010).

48. Saykin, A. J. et al. Alzheimer’s Disease Neuroimaging Initiative biomarkers as quantitative phenotypes: Genetics core aims, progress, and plans. Alzheimer’s and Dementia 6, 265–273 (2010).

49. Hooshmand, K. et al. Human Cerebrospinal Fluid Sample Preparation and Annotation for Integrated Lipidomics and Metabolomics Profiling Studies. Mol. Neurobiol. 61, 2021–2032 (2024).

50. Farr, E. et al. MetalinksDB: a flexible and contextualizable resource of metabolite-protein interactions. Brief. Bioinform. 25, (2024).

51. Szklarczyk, D., et al. The STRING database in 2023: protein-protein association networks and functional enrichment analyses for any sequenced genome of interest. Nucleic Acids Res. 51, D638–D646 (2023).

