## SupplementaryFigures for "Polyol pathway dysregulation in CSF links glucose metabolism to tau pathology independently of amyloid and genetic predisposition"

### Extended data

#### Extended Data Figure 1

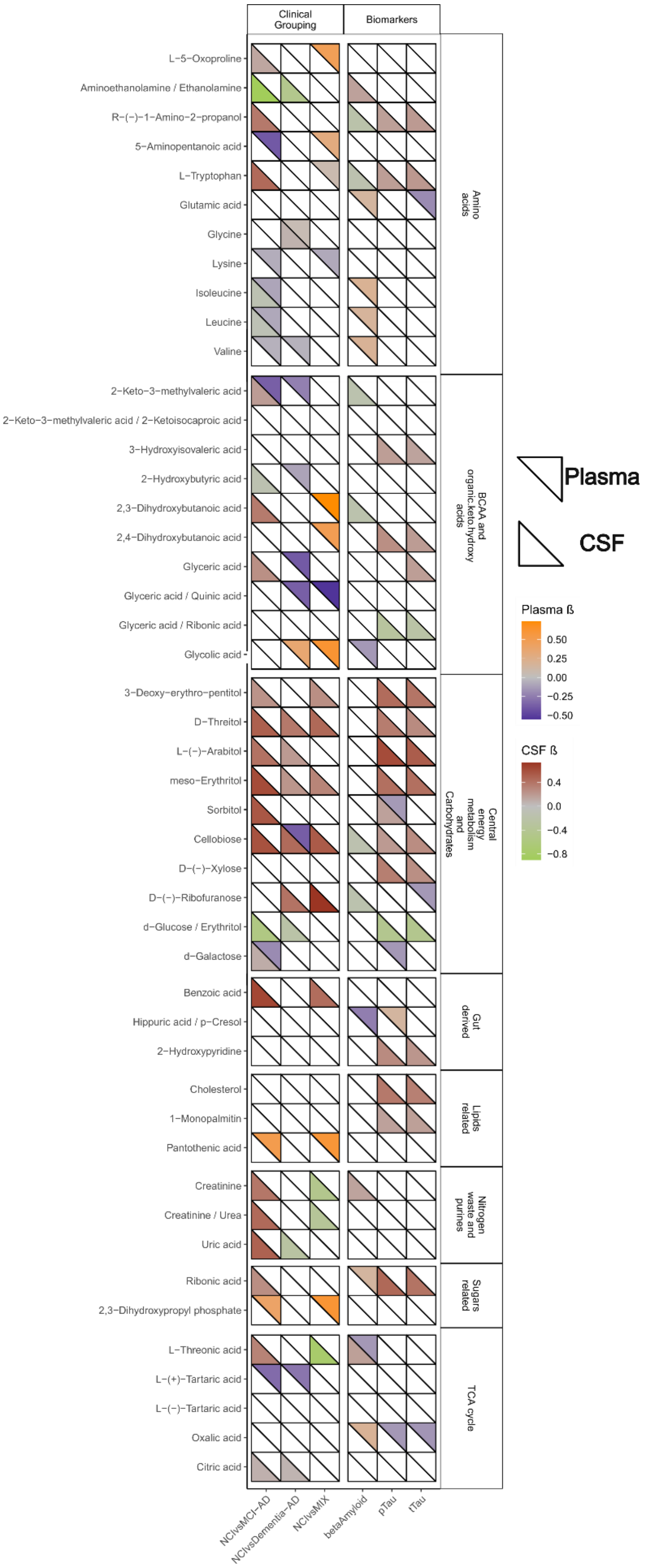
Clinical Grouping
Biomarkers
L−5−Oxoproline
Aminoethanolamine / Ethanolamine
R−(−)−1−Amino−2−propanol
5−Aminopentanoic acid
L−Tryptophan
Glutamic acid
Glycine
Lysine
Isoleucine
Leucine
Valine
Amino acids
2−Keto−3−methylvaleric acid
2−Keto−3−methylvaleric acid / 2−Ketoisocaproic acid
3−Hydroxyisovaleric acid
2−Hydroxybutyric acid
2,3−Dihydroxybutanoic acid
2,4−Dihydroxybutanoic acid
Glyceric acid
Glyceric acid / Quinic acid
Glyceric acid / Ribonic acid
Glycolic acid
BCAA and organic.keto.hydroxy acids
3−Deoxy−erythro−pentitol
D−Threitol
L−(−)−Arabitol
meso−Erythritol
Sorbitol
Cellobiose
D−(−)−Xylose
D−(−)−Ribofuranose
d−Glucose / Erythritol
d−Galactose
Central energy metabolism and Carbohydrates
Benzoic acid
Hippuric acid / p−Cresol
2−Hydroxypyridine
Gut derived
Cholesterol
1−Monopalmitin
Pantothenic acid
Lipids related
Creatinine
Creatinine / Urea
Uric acid
Nitrogen waste and purines
Ribonic acid
2,3−Dihydroxypropyl phosphate
Sugars related
L−Threonic acid
L−(+)−Tartaric acid
L−(−)−Tartaric acid
Oxalic acid
Citric acid
TCA cycle
NCIvsMCI-AD
NCIvsDementia-AD
NCIvsMIX
betaAmyloid
pTau
tTau
Plasma
CSF
Plasma ß
0.50
0.25
0.00
−0.25
−0.50
CSF ß
0.4
0.0
−0.4
−0.8

3

**Extended Data Figure 1: Associations between plasma and CSF metabolites and the clinical dementia categories and common biomarkers, filtered by multi-associated metabolites, either to both tissues or multiple categories or biomarkers.** Metabolite associations to tissues are represented in triangles, with the upper triangles in the tiles representing the plasma-associated metabolites and the lower triangles the CSF-associated metabolites. Associations to clinical categories are indicated with the mean beta coefficient, while associations to clinical biomarkers are indicated with the $r^2$ value. Only associations that were found to be significative ($p$-value $< 0.05$) were included in the figure. The heatmap has been split in blocks according to a major classification of metabolites, from top to bottom: sugar related metabolites; gut derived metabolites; nitrogen waste and purine related metabolites; lipids and related metabolites; branched-chain amino acids and organic/keto/hydroxy acids; amino acids and related ones; TCA cycle related metabolites; and central energy metabolism related metabolites.

### Extended Data Figure S2

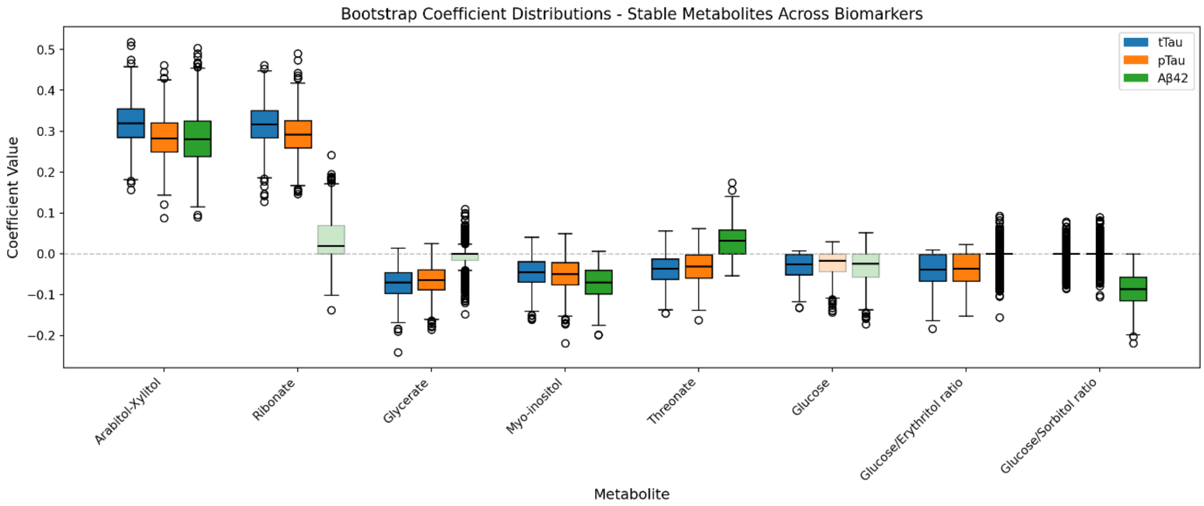


**Extended Data Figure 2:** Contributions of the tested polyol-related metabolites in the ElasticNet models for each of the three dementia biomarkers, as determined by the boxplot filling, across the bootstrap runs in the validation cohort. Stable metabolites (>70% of selection frequency) are opaque filled, while non-stable metabolites are filled with transparency.

### Extended Data Figure S3

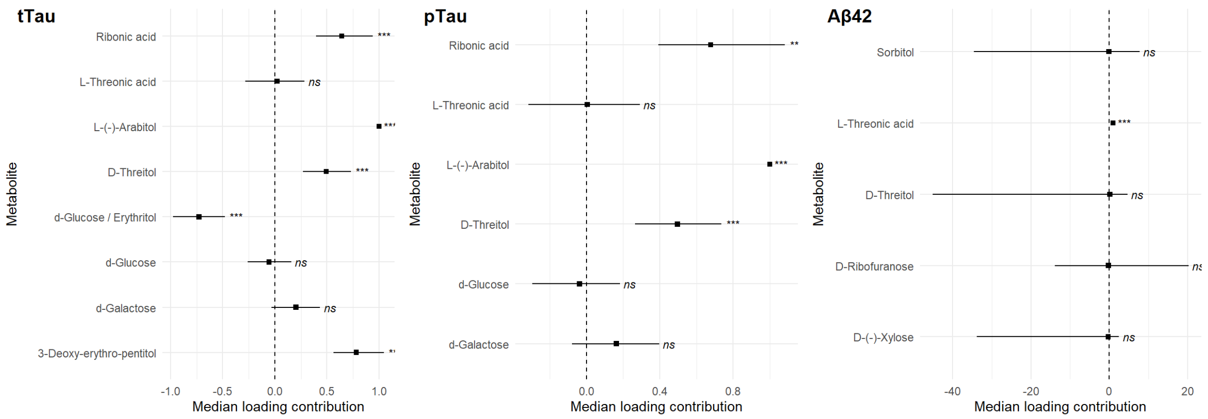


**Extended Data Figure 3:** Metabolite loadings for the SEM models in the NCI *vs* Dementia-AD contrast for the clinical biomarkers tTau, pTau and Aβ42. Points represent the median loading contribution of the 1,000 bootstrapping runs, while the horizontal lines cover the 95% interval of confidence.

### Extended Data Figure S4

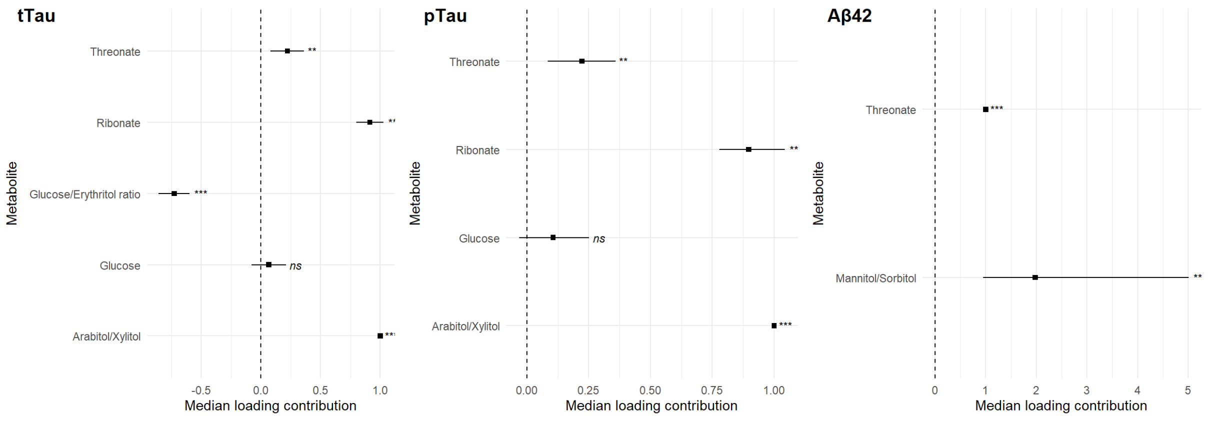


**Extended Data Figure 4:** Metabolite loadings for the SEM models in the NCI *vs* Dementia-AD contrast for the clinical biomarkers tTau, pTau and Aβ42 in the discovery cohort. Points represent the median loading contribution of the 1,000 bootstrapping runs, while the horizontal lines cover the 95% interval of confidence.
